# Impact on childhood mortality of interventions to improve drinking water, sanitation and hygiene (WASH) to households: systematic review and meta-analysis

**DOI:** 10.1101/2023.03.13.23287185

**Authors:** Hugh Sharma Waddington, Edoardo Masset, Sarah Bick, Sandy Cairncross

**Affiliations:** Environmental Health Group, Department of Disease Control, London School of Hygiene and Tropical Medicine (LSHTM), London International Development Centre (LIDC), 20 Bloomsbury Square, London, WC1A 2NS, United Kingdom; Department of Public Health, Environments and Society, LSHTM, and Deputy Director, Centre of Excellence for Development Impact and Learning (CEDIL), LIDC, 20 Bloomsbury Square, London, WC1A 2NS, United Kingdom; Environmental Health Group, Department of Disease Control, LSHTM, Keppel Street, London, WC1E 7HT

## Abstract

**Background:** In low-and middle-income countries (L&MICs), the biggest contributing factors to the global burden of disease in childhood are deaths due to respiratory illness and diarrhoea, both of which are closely related to use of water, sanitation and hygiene (WASH) services. However, current estimates of the health impacts of WASH improvements use self-reported morbidity, which may fail to capture longer-term or more severe impacts. Moreover, reported mortality is thought to be less prone to bias. This study aimed to answer the question: what are the impacts of WASH intervention improvements on reported childhood mortality in L&MICs?

**Methods and findings:** We conducted a systematic review and meta-analysis, using a published protocol. Systematic searches of 11 academic databases and trial registries, plus organisational repositories, were undertaken to locate studies of WASH interventions which were published in peer review journals or other sources (e.g., organisational reports and working papers). Intervention trials of WASH improvements implemented under endemic disease conditions in L&MICs were eligible, from studies which reported findings at any time until March 2020. We used the participant flow data supplied in response to journal editors’ calls for greater transparency. Data were collected by two authors working independently.

We included evidence from 24 randomized and 11 non-randomized studies of WASH interventions from all global regions, incorporating 2,600 deaths. Effects of 48 WASH treatment arms were included in analysis. We critically appraised and synthesised evidence using meta-analysis to improve statistical power. We found WASH improvements are associated with a significant reduction of 17 percent in the odds of all-cause mortality in childhood (OR=0.83, 95%CI=0.74, 0.92, evidence from 38 interventions), and a significant reduction in diarrhoea mortality of 45 percent (OR=0.55, 95%CI=0.35, 0.84; 10 interventions).

Further analysis by WASH technology suggested interventions providing improved water in quantity to households were most consistently associated with reductions in all-cause mortality. Community-wide sanitation was most consistently associated with reductions in diarrhoea mortality. Around one-half of the included studies were assessed as being at ‘moderate risk of bias’ in attributing mortality in childhood to the WASH intervention, and no studies were found to be at ‘low risk of bias’. The review should be updated to incorporate additional published and unpublished participant flow data.

**Conclusions:** The findings are congruent with theories of infectious disease transmission. Washing with water presents a barrier to respiratory illness and diarrhoea, which are the two main components of all-cause mortality in childhood in L&MICs. Community-wide sanitation halts the spread of diarrhoea. We observed that evidence synthesis can provide new findings, going beyond the underlying data from trials to generate crucial insights for policy. Transparent reporting in trials creates opportunities for research synthesis to answer questions about mortality, which individual studies of interventions cannot be reliably designed to address.

**Author summary:** *Why was this study done?:*

- The biggest contributor to the global burden of infectious disease in childhood in developing countries is mortality due to respiratory and diarrhoeal infections, both of which are closely linked to deficient water, sanitation and hygiene (WASH) availability and use.
- Multiple systematic reviews and meta-analyses of WASH-related morbidity have been conducted, but there is a shortage of rigorous, systematic evidence on the effectiveness of WASH improvements in reducing mortality.

*What did the researchers do and find?:*

- We conducted a systematic review and meta-analysis of the impacts of WASH interventions on all-cause and diarrhoea-related mortality in L&MICs, incorporating evidence from 35 studies comprising 48 distinct WASH intervention arms.
- We found significant effects on all-cause mortality among children aged under 5 of interventions to improve the quantity of water available (34 percent reduction), hygiene promotion when water supplies were improved (29 percent reduction), and community-wide sanitation (21 percent reduction).
- We also found significant effects of WASH interventions on diarrhoea mortality among under-5s (45 percent reduction).

*What do these findings mean?:*

- Interventions to prevent water-related mortality in childhood in endemic circumstances provide adequate water supplies to households, enabling domestic hygiene, and safe excreta disposal in the household and community.
- Systematic reviews can provide new evidence for decision makers but the approach we present is reliant on trial authors and journals adhering to agreed standards of reporting.

## 1. Introduction

Diarrhoeal diseases and respiratory infections are thought to kill 4.1 million people each year [1,2]. Half of these deaths are of infants and young children aged less than 5 years old [3], around 1.2 million of whom live in circumstances without adequate drinking water, sanitation, and hygiene (WASH) in low-and middle-income countries (L&MICs) [4]. The global burden of disease (GBD) for communicable causes is weighted heavily by mortality in childhood, the two biggest single causes of which are diarrhoea and respiratory infection. Ninety percent of the total diarrhoea GBD and 99 percent of the total respiratory infection GBD is due to years of life lost (YLL) (S1 Annex Table A1).

Unfortunately, studies of the effects of WASH interventions on diarrhoea and other causes of mortality are beset by such ethical and logistical difficulties that, with very few exceptions (e.g., [5]) practically none were carried out until recently (e.g., [6–8]). For example, it could not be ethical to allow young children to die when life-saving oral rehydration solution (ORS) is widely available and affordable. As a result, and in accordance with the recommendation of the Grading of Recommendations, Assessment, Development and Evaluations (GRADE) procedure [9], the focus shifted from mortality to morbidity – mainly from diarrhoea – as a more accessible outcome.

GBD estimates of WASH-related mortality are presently calculated using estimated coefficients on diarrhoea morbidity impacts from systematic reviews and meta-analyses. Estimates vary widely (S1 Annex Table A2), suggesting great imprecision affecting our measurement of the gravity of the diarrhoea problem, globally or in any specific context. Of the 44 systematic reviews included in a recent WASH sector-wide interventions evidence map [10], half of which concerned effects of WASH provision on diarrhoea, none had synthesised the evidence on mortality in childhood. The most recent systematic evidence on WASH interventions and diarrhoeal illness was reported in the Lancet in July 2022 [11].

A common finding in existing reviews is that bundling WASH together does not produce additive effects in comparison with single water, sanitation or hygiene technologies [12]. One major reason for this finding is bias in reporting. For example, the most common method of collecting health outcomes data in impact evaluations of WASH interventions is through participant report [10]. However, data on reported illness have been shown to be biased in open (also called “unblinded”) trials [13–16]. Perhaps carers might misrepresent illness to minimise the time spent with enumerators when data are collected repeatedly over time [17,18]. Social desirability bias may also arise where participants are inadvertently induced to report favourably.

Briscoe et al. [19] highlighted how diarrhoeal illness becomes normalised among highly exposed groups over time which leads to underreporting, a problem we might expect to become worse when reporting is done by someone other than the patient, in this case the child’s carer. Or illness may be acknowledged differently by sex [20], where girls who complain about pain are less likely than boys to be pacified by their carers and therefore may not report it. In other words, we may not see additive effects of multiple WASH technologies provided together if bias in the reporting of disease outcomes, rather than diarrhoea epidemiology, is driving the findings.

The key advantage of randomised controlled trials (RCTs) over other methods is the clarity with which randomisation balances unobservable differences across groups in expectation, not in any single trial, but over multiple draws from the population [21]. Thus the “gold standard” for evidence on health impacts from these studies uses meta-analysis of findings from multiple studies [22]. However, meta-analysis can also magnify biases, because is harder to identify errors where they pervade the whole data set. Some approach is clearly needed to address reporting bias. Of great potential concern is publication bias, the phenomenon whereby trials are more likely to be published if they find significant effects, a factor that is made more likely when trials are funded by private manufacturers, as has been common in studies of water treatment (chlorine, water filters) and hygiene (soap) [23].

In this paper, we present a different approach to estimate the health effects of WASH improvements. There is a large number of trials of WASH interventions, sufficient numbers on which to estimate global effects on mortality, even when the individual studies themselves did not aim to do so. We conducted a systematic review of the effects of WASH interventions on child mortality in L&MIC contexts, drawing on a number of sources including losses to follow-up due to mortality as reported in participant flows. It is an established finding that study participants do not misreport death, even in open studies [15,16]. This might be because death of a child is a rare and salient event. The crucial advantage of this approach, therefore, is that reported mortality is less prone to bias.

We sought to answer four review questions:

1. What are the effects of interventions promoting improved water supply, water treatment and storage, sanitation and hygiene in L&MICs on all-cause mortality, and to what extent do these effects vary by contextual factors?
2. What are the effects of WASH interventions in L&MICs on diarrhoea mortality, and to what extent do they vary by contextual factors?
3. What are the predicted effects of WASH interventions at different baseline mortality levels?
4. To what extent are the findings robust to potential biases at the individual study and review levels?

## 2. Methods

### Search and selection of studies

This review was registered with Prospero under registration number CRD42020210694 and is reported following the Preferred Reporting Items for Systematic Reviews and Meta-Analyses guideline (S2 PRISMA 2020 Checklist). A full description of the procedures followed for searches, study inclusion, outcomes data collection, analysis and reporting is presented in the published protocol [24]. Searches for literature were done as part of an evidence and gap map [10]. Studies selected were published at any time until March 2020. Eleven academic databases and trial registries (e.g., Cochrane, Econlit, Medline, OpenTrials, Scholar, Web of Science) and sources of nonpeer reviewed literature including databases and organisational repositories were searched (e.g., 3ie Repositories, J-PAL, IRC International Water and Sanitation Center, UNICEF, the World Bank and the regional development banks). We used reference snowballing, including bibliographic backreferencing and forward citation tracking of studies and existing reviews. As a measure to reduce publication bias, studies published in any format were eligible, and searches done of repositories of this information. As a measure to avoid language bias, studies published in English, French, Spanish and Portuguese were included, and searches done of repositories of this information. A priority search algorithm based on machine learning was used in filtering studies at title and abstract stage using EPPI-reviewer software [25]. Selection of studies was done by two authors working independently.

Eligible studies were RCTs and non-randomised studies of interventions (NRSI) promoting access to or use of WASH technologies to households in L&MICs in endemic disease conditions. We included new or improved water supplies, drinking water treatment and storage, sanitation and hygiene technologies, including those enabling or promoting hand-washing at key times and other beneficial household practices (e.g., the washing of food, clothing and fomites). We excluded trial arms with a major non-WASH component (e.g., nutrition interventions). We classified WASH interventions according to the “main WASH” technology provided, which was either water supply, water treatment and storage, sanitation or hygiene technologies provided or promoted alone, or multiple combinations of WASH technologies. It was also possible to characterise interventions by whether they provided any improvements in water supply, water treatment, sanitation and/or hygiene alone or in combination with others, which we refer to as “any WASH”. This was due to problems in clearly identifying all the components of an intervention. For example, a debate among practitioners suggested that hand hygiene messaging is usually incorporated in CLTS [26].

Counterfactual conditions were categorised as “improved” or “unimproved” according to the WHO/UNICEF Joint Monitoring Programme (JMP) classification. Improved water supplies were defined where the majority of households in the sample used drinking water from an improved source (e.g., piped water to the household, a community standpipe or protected spring) within a 30-minute round-trip including waiting time. For sanitation, the counterfactual scenario was defined as “improved” if the majority of controls had a sewer connection to the home or an improved pit latrine was used by a single household. Where insufficient information was reported about the counterfactual scenario to categorise baseline water supply or sanitation use, the figures were imputed from online data provided by the JMP for the relevant country, year and location.

A risk-of-bias tool was developed for WASH impact evaluations that drew on Cochrane’s tools for RCTs [27], cluster-RCTs [28] and non-randomised studies of interventions [29], and a tool for appraising quasi-experiments [30]. Six bias domains were assessed: confounding, selection bias, departures from intended interventions, missing data, outcome measurement bias and reporting bias. The studies were assessed on the likelihood of bias in estimating effects of WASH access on mortality in children aged under 5 years. This may or may not have been a primary research question in the papers themselves, hence our ratings do not provide risk-of-bias assessments for the study overall. The risk-of-bias assessments were done by two researchers working independently, at the outcome level for each included study arm, as recommended by Cochrane [22] and the Campbell Collaboration [31]. Template data collection forms are available in the study protocol [24]. Data extracted from included studies is provided in S1 Annex Table A3. The dataset used in analysis is provided in S4 Dataset.

### Measuring mortality outcomes

The primary outcomes for the review were all-cause mortality and mortality due to diarrhoeal disease. Outcomes data were collected independently by two researchers from two sources. The first source was the few studies that reported mortality alongside statistical information [6–8,32,33]. Mortality data were also recoverable from studies that reported losses to follow-up (attrition) in sample populations. Participant flow diagrams were reviewed in all studies of WASH technologies in L&MICs to obtain crude mortality rates for field trials by intervention group. These studies therefore formed the major source of evidence on all-cause mortality. Some studies also reported cause-specific mortality rates, including diarrhoea and other infections, defined by carers in verbal autopsy and/or clinicians, or collected from vital registries. All-cause mortality and mortality due to diarrhoea or other infections were defined by carers in self-report or taken from vital registries.

Mortality rates were computed over a standard period, as mortality measurements increase over longer exposure periods. Age-specific (e.g., under-2) mortality rates were defined where these data were available [6–8,34], or, if they were not, crude mortality rates were taken over the data collection period. Intervention effects were measured as the odds ratio (OR) of the mortality rates, and their 95 percent confidence intervals. Where studies reported multiple intervention arms against a single control arm, we split the control sample assuming an equal mortality rate for each comparison. We applied a continuity correction in study arms where there were no deaths, by adding 0.5 to all frequencies, which can cause bias in meta-analysis of rare events [35]. These studies were assessed as being at ‘high risk of bias’ in the outcome measurement domain [36–40].

### Evidence synthesis approach

Overall pooled effects were estimated for all-cause mortality (review question 1) and diarrhoea mortality (review question 2) using Stata. We assessed the consistency of the pooled effects using I-squared and tau-squared statistics to measure the relative and absolute heterogeneity between studies. We tested for effect moderators in meta-analysis and meta-regression analysis, including the WASH intervention technology provided to study participants, water supply and sanitation conditions in the counterfactual group, participant characteristics (age and if from immunocompromised group), and study characteristics (season of data collection and length of follow-up). We report forest plots showing WASH technologies for each analysis (we also report the same forest plots by study author in S3 Annex Figs A1-A4). To aid interpretation of the meta-regression coefficients, we calculated OR prediction values at the means, minima and maxima of the dichotomous variables and the mean and interquartile range of the continuous variable. Moderator variables were pre-specified based on theory and previous reviews, with the exception of the moderator analysis by baseline mortality rate. We used meta-regression plots to assess the predicted effects of the interventions by baseline mortality rate (review question 3).

We evaluated the likelihood that potential biases could cast doubt on the findings results through a negative control, formal publication bias assessment, and sensitivity analysis (review question 4). The effects of WASH improvements on mortality are largely expected to occur by blocking transmission of infectious diseases, primarily faeco-oral and respiratory infections, in childhood. People who survive beyond the age of 5 are thought to have developed sufficiently robust immunity to these diseases, hence the effects of WASH improvements on mortality among older groups is expected to be far weaker. Therefore, as a negative control [41,42], meta-analysis was estimated for those studies that reported all-cause mortality among participants aged over 5 years. We also assessed the sensitivity of the pooled effects to exclusion of each single effect, examined whether there was a correlation between risk-of-bias rating and the estimated effect, and tested for small-study effects (publication bias) at the review level using graphical inspection of funnel plots and regression tests.

## 3. Results

### Description of searches and included studies

From 13,500 de-duplicated records, 684 full text reports of WASH intervention studies were screened, of which 35 were identified that reported mortality outcomes, 30 of which were measured among children aged 5 or under (Fig 1). We were not able to incorporate trials that met the review inclusion criteria but did not report participant flows (e.g., [43]). We found 24 RCTs that measured mortality, all of which were published in peer review journals. RCTs were of water treatment and storage, sanitation and/or hygiene interventions, which mainly used cluster design, with clustering at the community level. We found no RCTs of water supply provision or promotion that reported mortality estimates. Several studies used prospective non-randomised trial designs [33,36,44], five analysed cohort data [38,45–48], one used a matched pipeline approach [49] and two used repeated cross-section data with double-differences [50,51]. Six of the studies were designed retrospectively after the WASH intervention had been conducted [47–51]. The effect of the WASH intervention was not calculable for one non-randomised study [51]. All RCTs were reported in English. Of the nine included non-randomised studies of interventions, which were published in peer review journals and reports, three were in French [33], Spanish [49] or Portuguese [44]. The studies were published from 1985 onwards, the majority in the 2010s. The evidence is representative of all lower-income global regions and many relevant contexts, including rural, urban and peri-urban informal settlements.

**Fig 1.**
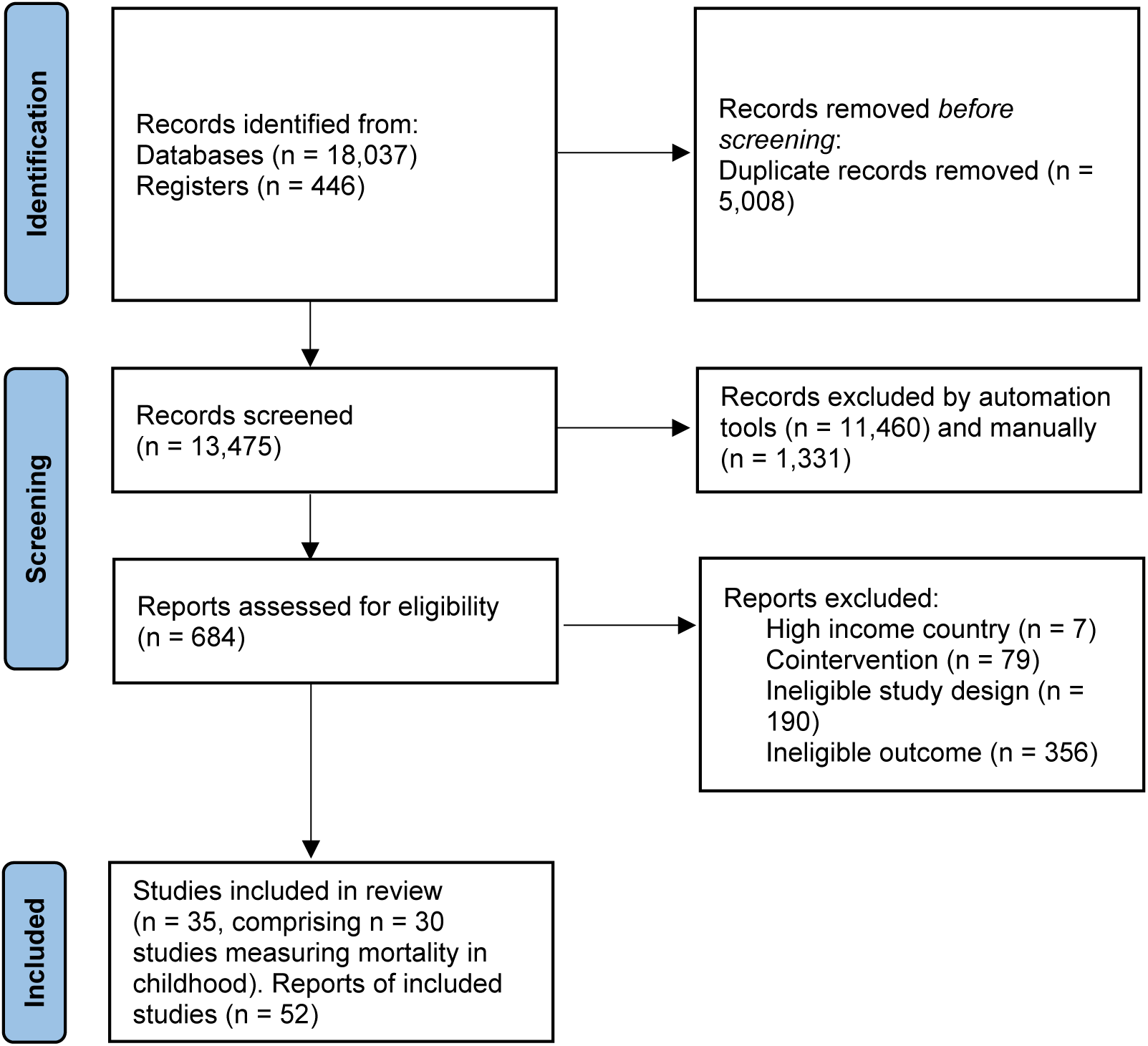
PRISMA study search flow.

We included in meta-analysis 38 WASH study arms examining all-cause mortality in childhood, of which 26 were from RCTs, and 10 examining diarrhoea mortality, of which 6 were from RCTs. For six studies we could also extract seven estimates of effects on mortality among adults and/or children aged over-5 [32,39,47,52–54]. In five studies comprising seven study arms, mortality was reported for all age groups combined [39,40,55–57]. There was a total of 168,500 participants in the included studies of all-cause mortality and 2,600 deaths. When including the natural experiment of Galiani et al. [50], we estimated there were 165,000 more child deaths.

We grouped the interventions by WASH technology provided. Many concerned direct hardware provision – water supplies, filters, handwashing stations and/or latrines – and health messaging. Thus, the WASH technology provided in six studies was household water treatment by chlorine alone [6,8,37,40,54,57] or alongside safe storage devices [56,58]. Three studies evaluated filter provision with safe storage [59–61] and two evaluated UV irradiation (solar disinfection, SODIS) [62,63]. A further 11 studies incorporated arms evaluating hygiene promotion alone [6,8,34,36,39,40,45,58,64–66]. Others included arms combining household water treatment with handwashing promotion [38,40,65] or alongside handwashing and sanitation [6–8]. A water supply improvement was provided alone in three non-randomised studies [46,48,67], another concerned improved water supply and sewage connections [50], three were of water supplies and latrines [44,47,49], and one other was of water supply, latrines and handwashing promotion [33]. Three study arms were evaluated of latrine provision or promotion alone [8,8,52], but ten studies evaluated sanitation alongside other WASH technology improvements [6–8,32,33,44,47,49,50,53]. For example, the Total Sanitation Campaign in India provided hygiene education alongside CLTS, subsidies and sanitation marketing [53].

Counterfactual groups often received standard WASH access with no additional interventions, although occasionally they received another intervention; for example, all participants received hygiene education in one study [56]. Most counterfactual populations were assessed as using improved water supplies [7,8,8,34,36–38,40,44,45,48–50,53,57,58,63–66]. In a few instances, counterfactuals received piped water inside the compound [36,50], otherwise it was sourced by household members from outside. In one study of continuous water supply (“safely managed drinking water”) provision, controls received water for only a few hours a week on average [48]. There were also concerns about reliability of or distance to the water supply in a few studies [8,65], which would have affected ability of study participants to practice improved hygiene. In under half of cases, sanitation was classified as being improved [6,36,38,40,44,48–50,56,63–65]. In all others, the majority of households openly defaecated, or used shared facilities or unimproved facilities like pits without concrete slabs. Imputations were made where it was not clear exactly what types of water and sanitation services were used by households in the counterfactual scenario [33,34,37,38,44–46,49,50,52,55,58,61].

### Assessment of biases at the study level

In general, just under half of studies (40%) were found to be at ‘moderate risk of bias’ overall in attributing changes to the intervention, for all-cause mortality (Fig 2a) and mortality due to diarrhoea (Fig 2b). No studies were at ‘low risk of bias’.

**Fig 2.**
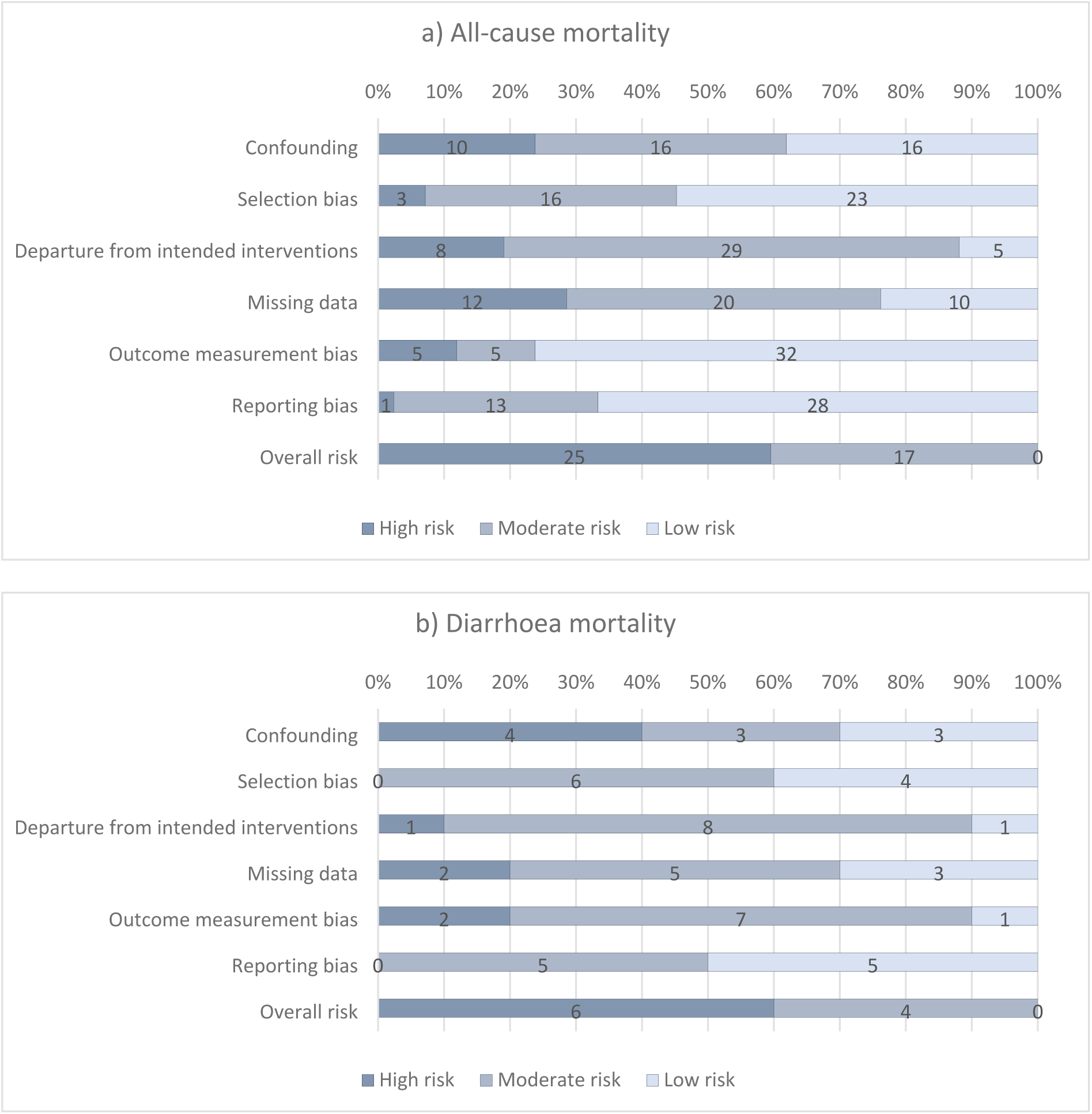
Risk-of-bias assessments.

One-third of RCTs reported using adequate allocation sequence and concealment, and demonstrated baseline covariate balance, to satisfy a ‘low risk’ rating on confounding. In some cases, data were collected on water, sanitation and hygiene at pre-test, but balance was not presented for all relevant variables, such as sanitation and hygiene access. Three NRSI were assessed as being at ‘moderate risk of bias’ in confounding. These were all studies of water supply improvements including: privatised water provision in Argentinean municipalities [50], improved water supply reliability in India [48], and piped water supply and latrines in India [47]. In all cases, participation was largely determined by programme placement, which is thought less problematic to address than self-selection into programmes by participants. In Argentina, it was the local government’s decision to implement a central government policy allowing for privatisation of the water supply [50]. For piped water in India, all households in a community were simultaneously connected to the water supply by the NGO Gram Vikas [47]. For the study examining the reliability of water supplies in India, all households were connected to the municipal supply [58]. Participation was then carefully modelled using a rich set of covariates measured at baseline and based on factors thought to influence programme targeting. Each study also presented null findings for a negative control (placebo outcome): mortality due to non-infectious causes [50], and the incidence of bruising and scrapes [47,48].

Where participants were recruited before allocation in cluster-RCTs, or where recruiters were blinded to allocation, the studies were judged to be at ‘low risk’ of selection bias. Where recruitment was done afterwards by those potentially with knowledge of allocation or where individuals needed to be recruited later due to attrition (losses to follow-up during the trial), the study was judged to be at risk of bias. Studies were also assessed as being at ‘high risk of bias’ when overall attrition rates were greater than 20 percent, or differential attrition greater than 10 percentage points, or where no information was provided about reasons for dropouts by intervention group, tests for covariate balance or robustness of findings. Selection bias and attrition bias were deemed less problematic where studies used census data [50] or vital registration [44].

In general, departures from intended interventions due to contamination (controls receive the treatment) or spill-over effects (control outcomes are caused by treatment outcomes) were judged unlikely to be problematic in many studies, which used cluster-randomisation and reported geographical separation of groups. Of specific relevance to mortality estimates, studies providing ORS to severely ill children and/or encouraging mothers to attend health clinic were judged to have high risk of bias in the outcome measure.

Regarding outcome measurement, all-cause mortality was usually categorised as being a reliable measure even when self-reported with long recall, owing to the salience and rarity of the event; the longest recall was 6 years [65], the shortest two days [38], and usually it was 12 months or less. However, there is greater suspicion about cause-specific mortality where reporting is through verbal autopsy by the child’s carer. If cause-specific mortality was measured, assessment was therefore made as to whether it was verified by a clinician or taken from vital registration, in which case it was assessed as being at ‘low risk of bias’. While observational studies of WASH provision have verified cause of death through consultation with a clinician [5], no RCTs and only two NRSI used vital registration data [44,50]. One study [44] was assessed as at ‘low risk’ of outcome reporting bias for diarrhoea mortality, while another was assessed as at ‘high risk of bias’ because the study did not attribute cause-specific mortality to diarrhoea, using infectious and parasitic disease mortality instead [50]. In all other studies the cause of death was given by verbal autopsy.

Nearly all trials were pre-registered, four reported publishing a protocol with pre-analysis plan [6–8,60], and three blinded data analysts [6–8]. In addition, one NRSI was deemed to have ‘low risk of bias’ on reporting, because it published a baseline report with pre-analysis plan [68].

### Impacts of WASH on all-cause mortality (review question 1)

We conducted meta-analysis across intervention arms reporting all-cause mortality in children aged under 5 years (Fig 3). WASH improvements typically reduced the odds of all-cause mortality in childhood by 17 percent overall (OR=0.83, 95%CI=0.74, 0.92, 38 estimates). There was a small degree of estimated relative heterogeneity (I-squared=16%) and absolute heterogeneity (tau-squared=0.01).

**Fig 3.**
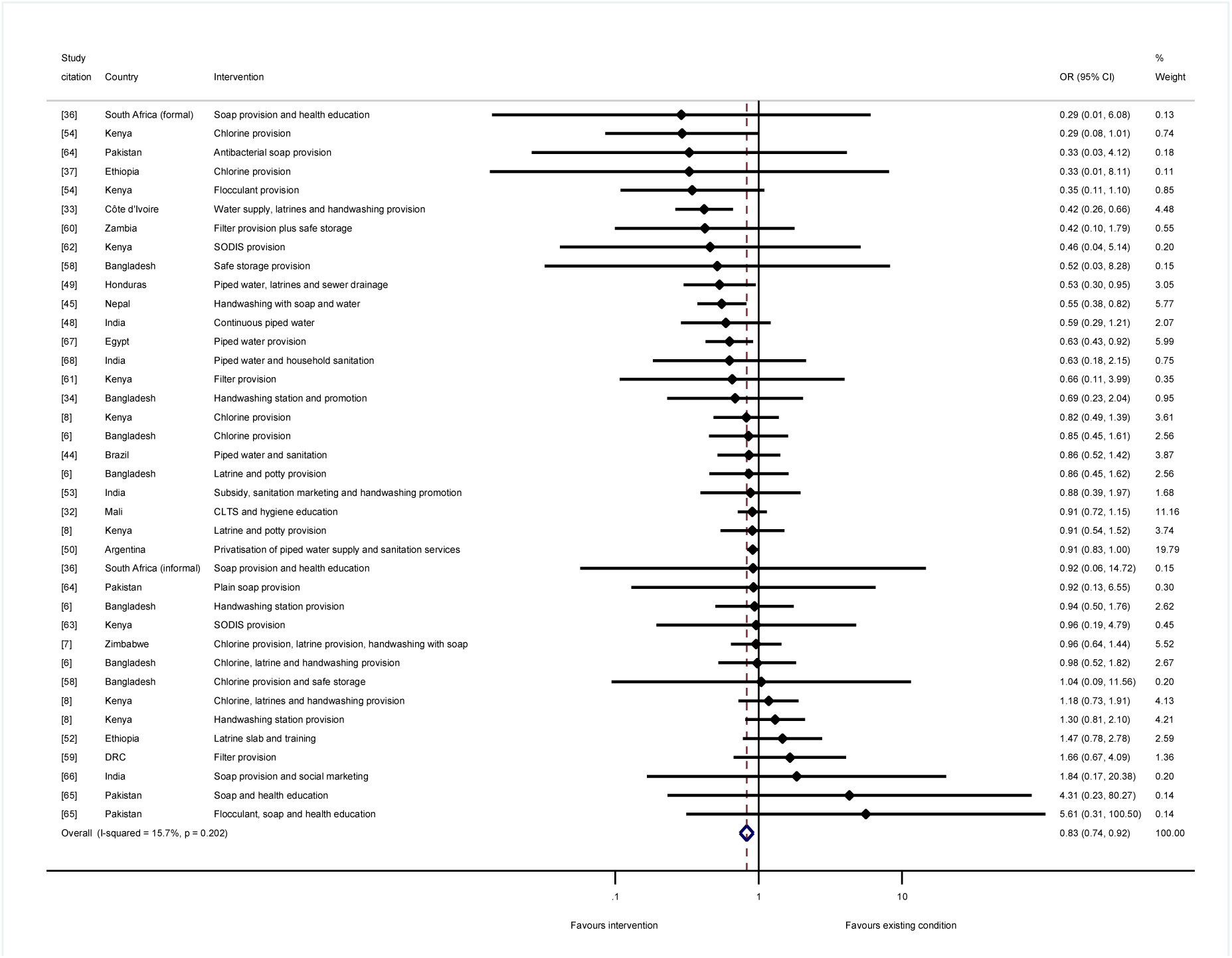
Effects on all-cause mortality in childhood of WASH improvements.

For the stratified meta-analyses by WASH technology, trial arms incorporating “any WASH” – that is, any single water supply, water treatment, sanitation or hygiene technology, whether provided alone or alongside any other WASH technology – were meta-analysed. We found a 34 percent reduction in the odds of mortality for water supply improvements (OR=0.66, 95%CI=0.50, 0.88; I-squared=66%; 7 estimates) (Fig 4a). Four of these were studies where the risk of bias was high [33,44,49,67], while three were at ‘moderate risk of bias’ [47,48,50]. For sanitation, we estimated 13 percent reduction in mortality overall (OR=0.87, 95%CI=0.75, 1.00; I-squared=33%; 13 estimates). Four of the studies were assessed as being at ‘high risk of bias’ [33,44,49,52], and seven were at ‘moderate risk of bias’. We tested for a threshold effect of sanitation improvement – that is, whether there needed to be a certain share of households in a community covered before the benefits of sanitation were realised [69]. When sanitation improvements targeted the whole community rather than individual households, or if households were targeted for sanitation improvements in circumstances when most of the community already used improved sanitation facilities, there were greater effects on mortality among children participating in the study (Fig 4b). There was an estimated 21 percent reduction in the odds of mortality when sanitation was being improved community-wide (OR=0.79, 95%CI=0.66, 0.95; I-squared=43%; 8 estimates), but no effect of sanitation where it was provided to specific households, where the majority of community members did not already use improved sanitation (OR=1.07, 95%CI=0.83, 1.36; I-squared=0%; 4 estimates).

**Fig 4.**
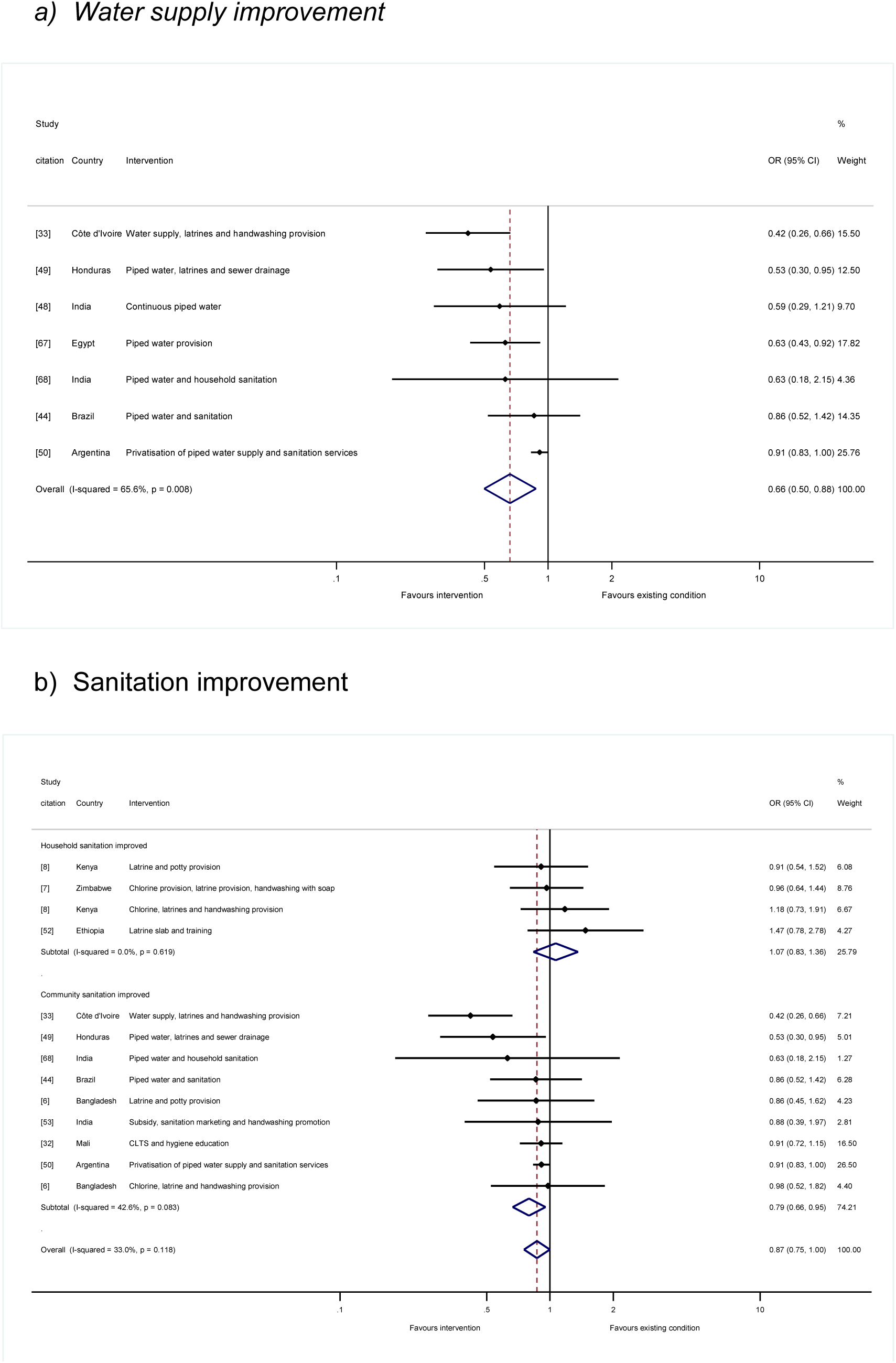
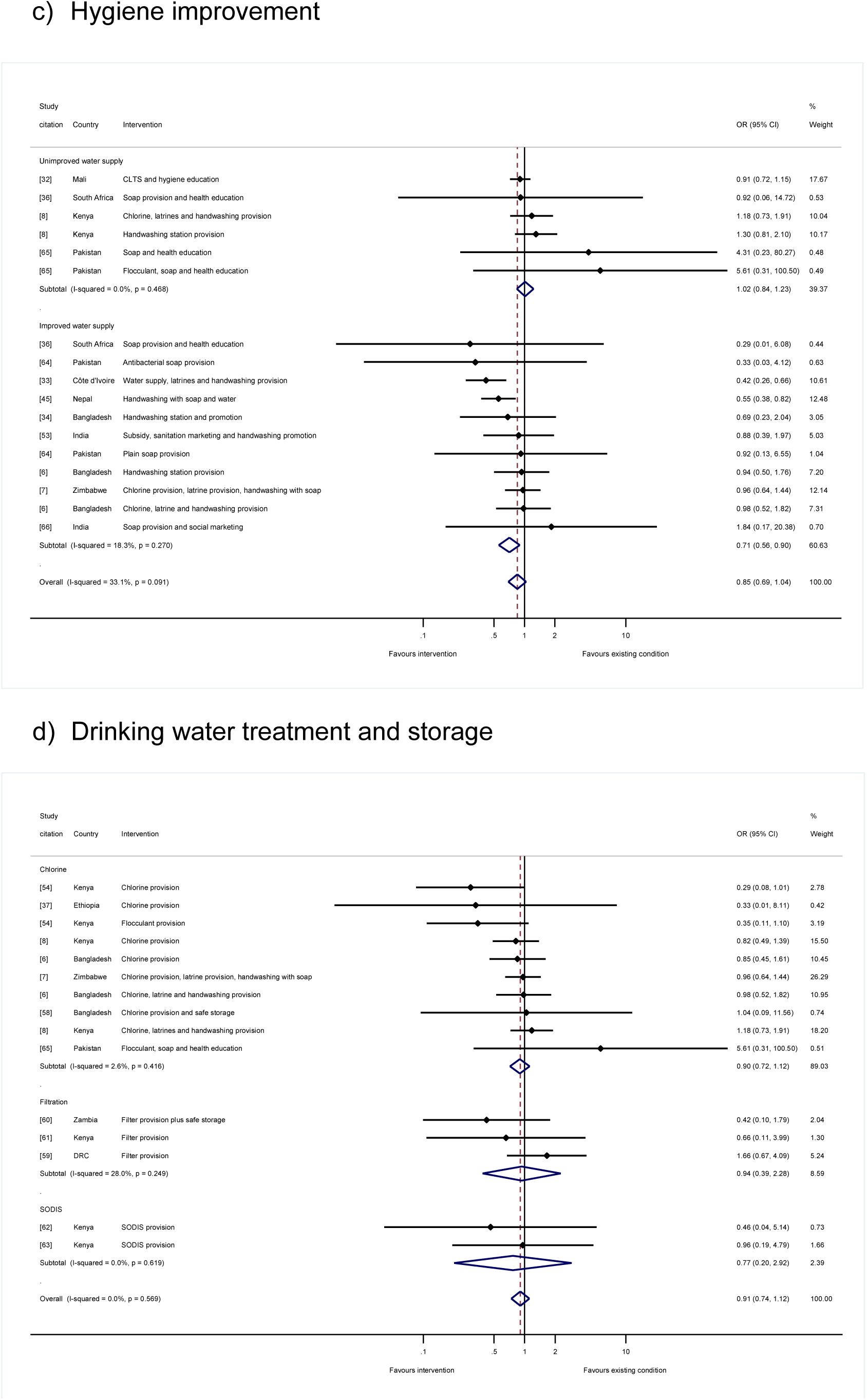
Effects on all-cause mortality in childhood by “any WASH” technology.

The overall effect of hygiene promotion was not statistically significant (OR=0.85, 95%CI=0.69, 1.04; I-squared=33%; 17 estimates). Five of the studies were assessed as being at ‘high risk of bias’ [33,36,45,65,66] and seven were at ‘moderate risk of bias’. Further analysis was done to test the hypothesis that hygiene promotion would be more effective when done under conditions of improved water supply, or, if not, when water supply was an intervention component alongside hygiene, and there were no concerns about the reliability of or distance to the water supply. The results suggested that this was indeed the case: there was no estimated effect of hygiene in circumstances where water supplies were not already improved (OR=1.02, 95%CI=0.84, 1.23; I-squared=0%; 6 estimates). In contrast, there was a 29 percent reduction in the odds of mortality, when hygiene was provided in circumstances where the water supply was also being improved or had been improved previously (OR=0.71, 95%CI=0.56, 0.90; I-squared=18%; 11 estimates) (Fig 4c).

There were no significant effects on mortality of household water treatment and storage overall (OR=0.93, 95%CI=0.75, 1.14; I-squared=0%; 15 estimates), or for individual water treatment technologies including chlorination (OR=0.90, 95%CI=0.72, 1.12; I-squared=2%; 10 estimates), filtration (OR=0.94, 95%CI=0.39, 2.28; I-squared=28%; 3 estimates) or SODIS (OR=0.77, 95%CI=0.20, 2.92; I-squared=0%; 2 estimates) (Fig 4d). Five of the studies were assessed as being at ‘high risk of bias’ [59,61–63,65] and seven were at ‘moderate risk of bias’.

Meta-analysis by the “main WASH” technology that was provided suggested reductions in odds of death in childhood which were of the same magnitude but not statistically significant for any single technology provided alone. But there was a significant reduction in mortality where multiple water, sanitation and/or hygiene technologies were promoted or provided of 16 percent (OR=0.84, 95%CI=0.71, 0.99, I-squared=41%, 11 estimates). Five of the seven studies with the largest effects of multiple WASH technologies incorporated a water supply improvement [33,44,47,49,50], usually piped water to the household or yard.

We estimated meta-regressions to explore further whether the variation in effects by WASH technology intervention, and the other contextual factors we had identified from theory, might explain differences across studies (Table 1). The regression pooled data from study participants of any age, incorporating the 14 additional estimates measured among all population groups or adults and children aged over 5. The reductions in mortality were significantly larger when interventions were conducted in circumstances where: participants were children aged under 5 years, or data collection was limited to the summer rainy season. Where the study collected data over a shorter follow-up period, the effect on mortality was also significantly larger. Impacts on mortality were significantly greater when water supply improvements were made. The explanatory power of the regression was high (R-squared=76%) and there was very little residual heterogeneity (I-squared=0%; Tau-squared<0.01). The findings suggested a predicted value of 12 percent reduction in odds of mortality at the data means (OR=0.88) (Table 1 Panel 3). The maximum value of 74 percent reduction in odds of mortality (OR=0.26) is for children from immunocompromised groups who would receive all WASH interventions, with measurement made against comparators living in very poor communities with unimproved sanitation services, during the summer rainy season at 6-months intervention follow-up.

**Table 1.**
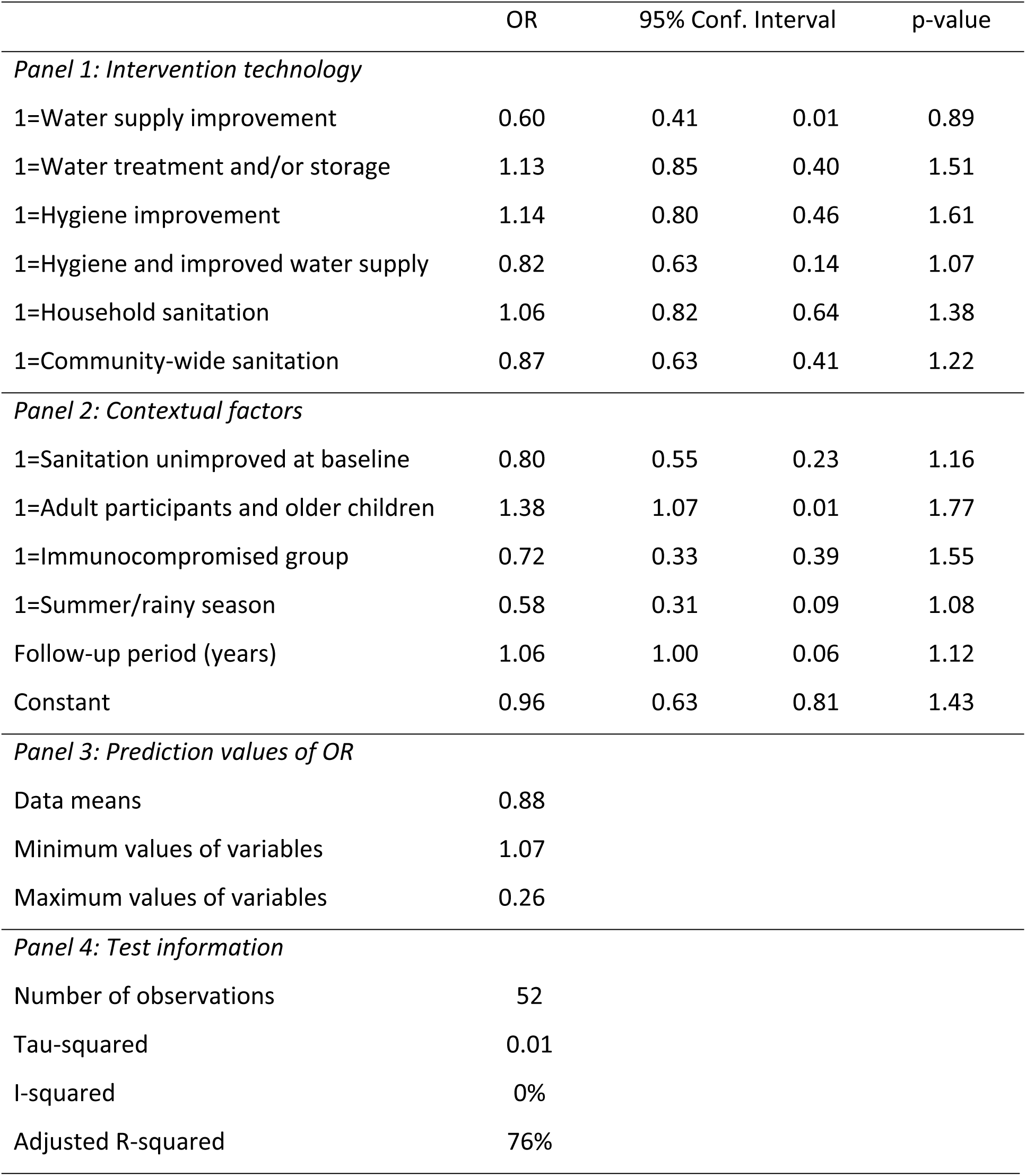
Meta-regression analysis of all-cause mortality with prediction values

### Impacts of WASH on diarrhoea mortality (review question 2)

The meta-analysis of diarrhoea mortality in childhood suggested WASH provision and promotion lead to a reduction in the odds of death due to diarrhoea by 45 percent (OR=0.55, 95%CI=0.35, 0.84; 10 estimates) (Fig 5). Six of the studies were assessed as being at ‘high risk of bias’ [33,38,44,46,50,65] and three were at ‘moderate risk’ [32,60,64]. The relatively high degree of absolute and relative heterogeneity in findings (I-squared=43%, tau-squared=0.15) suggested additional analysis was needed of factors that could explain the variation across study contexts.

**Fig 5.**
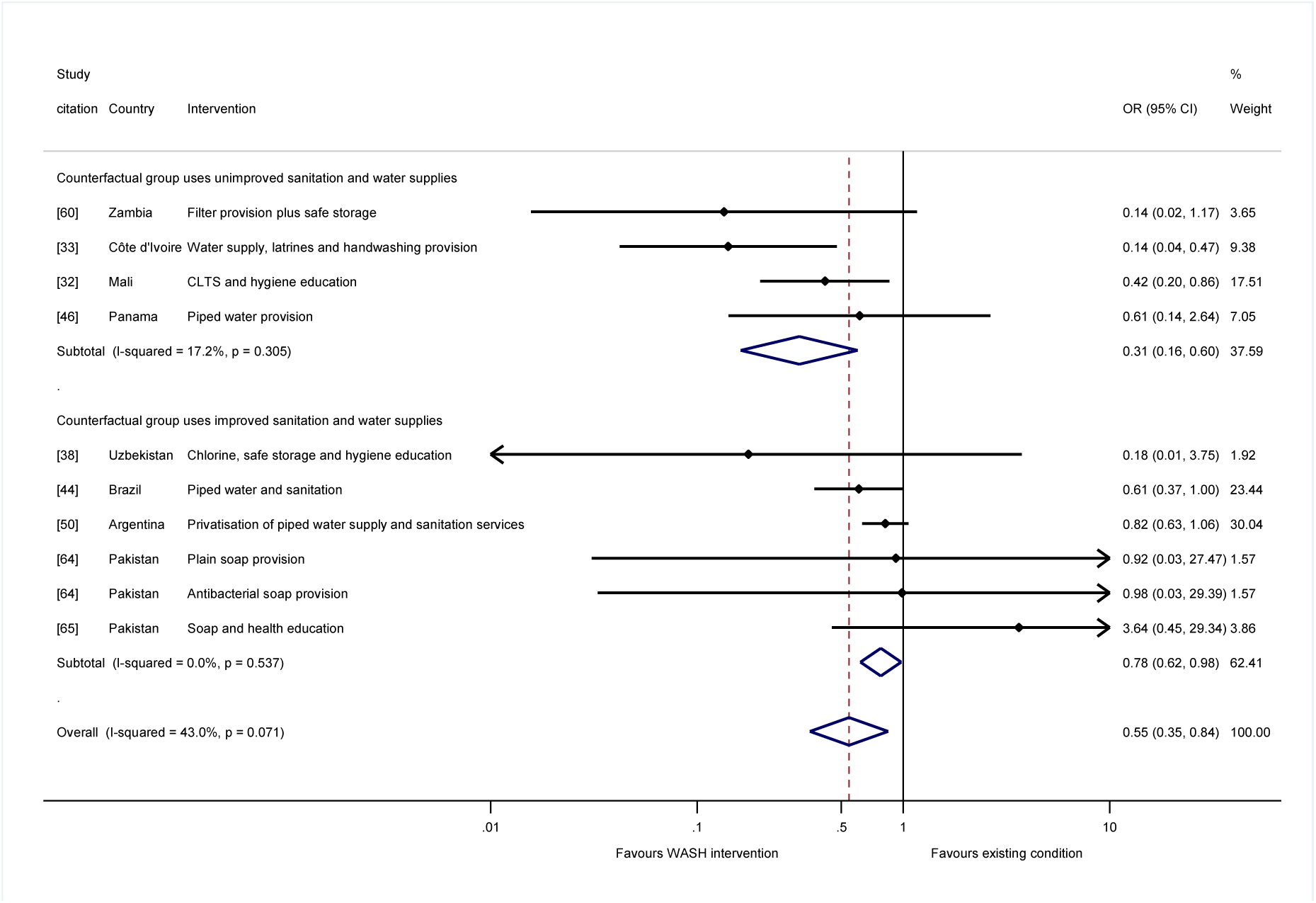
Effects on diarrhoea mortality in childhood of WASH improvements.

One of those factors is the degree of movement along the WASH ladders. We tested this hypothesis in moderator analysis according to the type of water supply and sanitation facilities used in the counterfactual group. When the WASH interventions were provided when counterfactuals were using no or unimproved sanitation and water supplies, and therefore exposed to very high risk of environmental contamination by pathogens, there was an estimated 69 percent reduction in diarrhoea mortality in childhood (OR=0.31, 95%CI=0.16, 0.60, I-squared=17%, 4 estimates). But for interventions provided in circumstances when most people already had access to improved water supply and sanitation, there was only a 22 percent reduction in odds of mortality (OR=0.78, 95%CI=0.62, 0.98, I-squared=0%, 6 estimates) (Fig 5). The impacts of WASH interventions on childhood diarrhoea mortality were significantly greater (p<0.01) when counterfactual groups lacked access to improved water supply and sanitation – and most people were therefore using unimproved facilities, or none at all and openly defaecating – than when most people in counterfactual groups were using improved facilities.

The largest effects on diarrhoea mortality were from studies of multiple WASH technologies: two contained a component that aimed to provide latrines to all households in intervention communities [32,33] and two involved water supply improvements [33] or hygiene promotion when water supplies were already improved [38]. With regard to the two studies of latrine provision or promotion to whole communities, both were provided alongside hygiene promotion, but only in Côte d’Ivoire was the water supply also improved [33]. In the case of the CLTS intervention in Mali [32], hygiene promotion was given when water supplies were limited. Another longitudinal follow-up study of an RCT of hygiene improvement, which was rated at ‘high risk of bias’, was conducted among communities where some households had access to running water for only two hours each week [65], suggesting these households had limited opportunities for adherence to improved hygiene practices.

Few studies of household water treatment in endemic circumstances have reported diarrhoea mortality outcomes. Among the studies examining HWT, only one was of an approach which has been found to reduce diarrhoea morbidity; the study was of filtration [60] and it found large but statistically insignificant impacts in children from immunocompromised populations (HIV-positive mothers). The other was a study of chlorine provision alongside safe storage and hygiene education [38]. Meta-regression analysis suggested interventions providing community-wide sanitation, and hygiene promotion in circumstances when water supplies were improved, were associated with significantly larger impacts on diarrhoea mortality (S1 Annex Table A4).

### Predicted effects of WASH improvements by baseline mortality rates (review question 3)

We tested for a theoretical relationship between the contextual starting values and programme effectiveness – that is, one might expect higher returns from a lower base – by plotting the relationships between the baseline mortality rate measured in the counterfactual group and the log-odds ratios for all-cause (Fig 6a) and diarrhoea mortality (Fig 6b). The results suggested that, at higher baseline mortality rates, WASH interventions tended to have larger effects on mortality. For example, where the crude mortality rate was 75 per 1,000 live children, as it is in many African countries and communities in South Asia, the estimated reduction in odds of all-cause mortality in childhood was 33 percent (OR=0.67, 95%CI=0.47, 0.86). At the same baseline mortality rate, there was a reduction of 61 percent in the odds of diarrhoea mortality (OR=0.39; 95%CI=0.20, 0.67).

**Fig 6.**
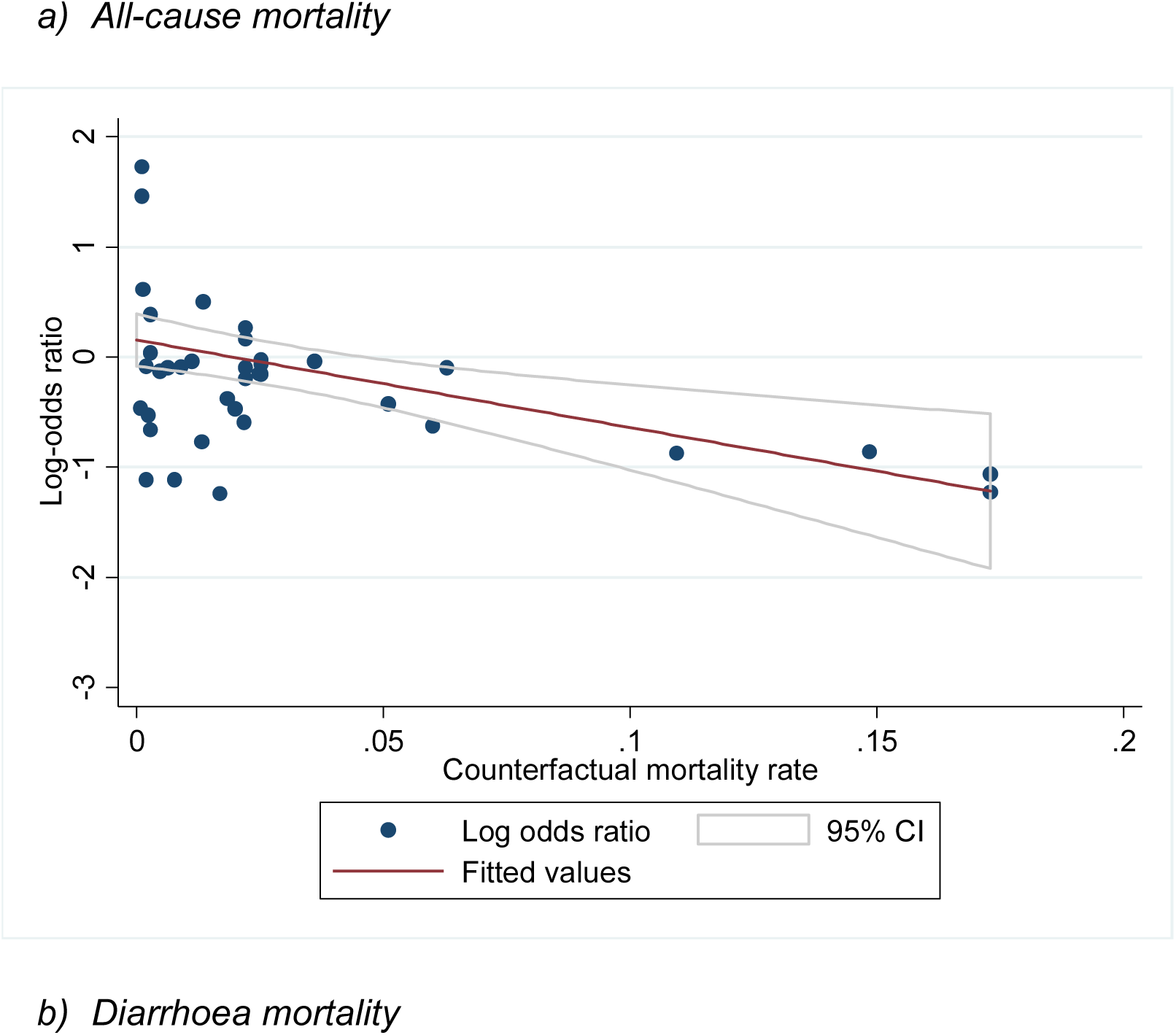
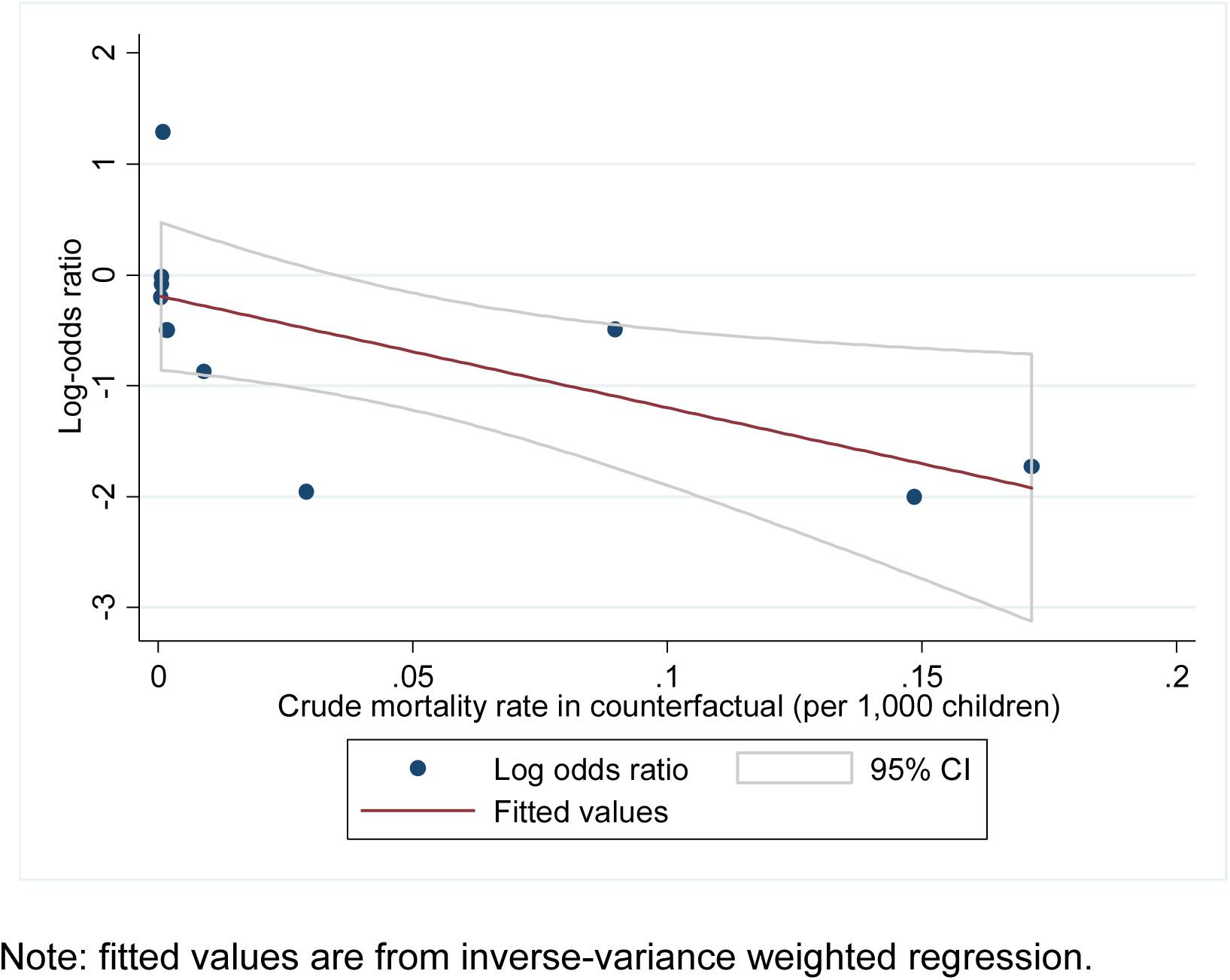
Meta-regression plots of all-cause mortality in childhood against prevalence.

### Evaluation of biases at the review level (review question 4)

In this section we present findings from a negative control, analysis of small study effects and the results of sensitivity analyses. Using meta-analysis to power studies adequately with small effect sizes does not necessarily generate effects that are statistically significant if there is no underlying causal relationship [70]. The meta-analysis of studies reporting all-cause mortality among participants aged over 5 years did not suggest WASH improvements affected mortality when participants were restricted to adults and children aged over 5 (OR=1.05, 95%CI=0.93, 1.19, I-squared=0%, 7 estimates) (Fig 7). The study with the largest effect on mortality was of health messaging among 10-year-old school children [39]. Several of the studies were of chlorination [54,56,57]. We might expect to see effects on maternal mortality due to sepsis, which improved WASH – particularly in places of birth like health facilities – is thought to alleviate [71]. None of the interventions provided a WASH intervention in a health facility.

**Fig 7.**
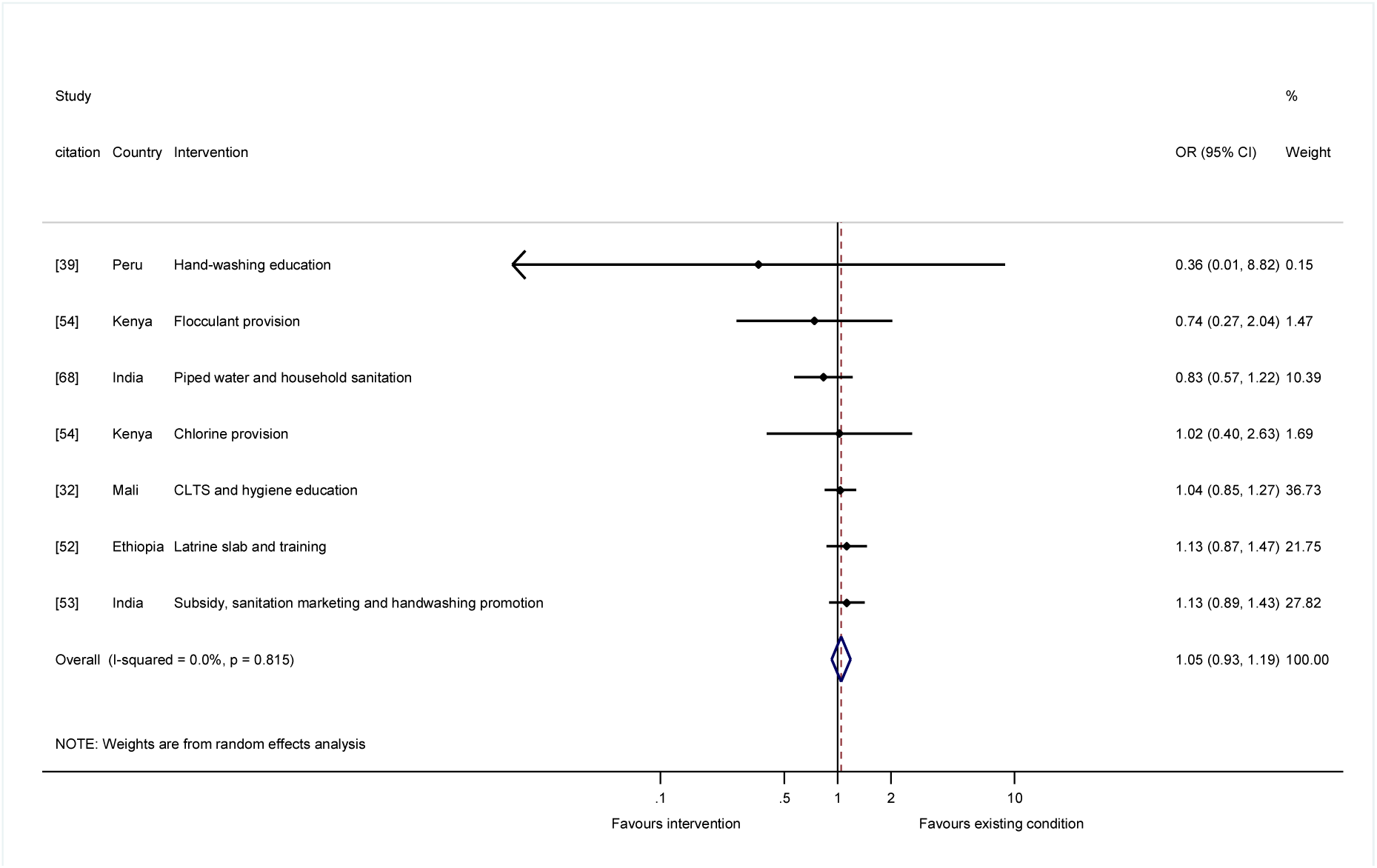
Effects on all-cause mortality for study participants aged over 5 years.

Since the mortality data were largely collected from participant flow diagrams, the fact that mortality estimates are available at all is indicative of the good standards of reporting in the studies included in this review. This suggested publication bias was likely to be limited, most clearly for prospective trials of WASH interventions, as found in the analysis of small study effects (S3 Annex Fig A5). We also tested the sensitivity of the findings to exclusion of particular studies. For example, the pooled effect estimate might be influenced by studies with large samples [50] or those conducted among extremely poor or vulnerable groups [32,60]. The overall findings, and the findings for particular WASH technologies or circumstances, were not significantly affected by exclusion of these, or any other individual studies. We also examined whether there was a correlation between risk-of-bias rating and the estimated effect on mortality. The effects assessed at ‘high risk of bias’ incorporated studies that did not distinguish under-5s from other population groups [40,57]. The meta-analysis of NRSI at ‘high risk of bias’ found a greater reduction in odds of all-cause mortality than other studies (OR=0.58, 95%CI=0.48, 0.70; I-squared=0%; 8 estimates) (S3 Annex Fig A6a). In contrast, we found no significant change in the odds of death for RCTs that had ‘high risk of bias’ in measuring the effect on mortality in children aged under 5 years (OR=1.41; 95%CI=0.99, 2.01; I-squared=0%; 15 estimates) (S3 Annex Fig A6b).

## 4. Discussion

### Summary of main findings

This systematic review and meta-analysis estimated the impacts of WASH improvements on children’s mortality by pooling data, collected mainly from reported participant flows in multiple studies, for all-cause and diarrhoea mortality. The approach helped overcome two critical issues in primary study research. Firstly, it is difficult to design prospective impact evaluations like RCTs with sufficient statistical power to estimate precise effects for rare outcomes. And, secondly, mortality is thought to be reported with less bias than other measures. The findings suggested WASH improvements cause large and statistically significant reductions in the odds of mortality in childhood in endemic circumstances. For mortality due to any cause, we estimated around one-in-five deaths are averted by WASH improvements. For severe diarrhoea disease we estimated a reduction in odds of mortality by nearly half. However, these averages concealed important heterogeneity in effects. Further analysis suggested that the reduction in all-cause mortality was most consistently established where the interventions provided an improved water supply.

Since many of the studies examining water improvements were of piped water to the household or yard, the analysis therefore suggested a mechanism through which water affects mortality: by enabling domestic hygienic practices around handwashing, food preparation and cleanliness. Indeed, where hygiene was promoted, the analysis suggested it was effective in circumstances where there was likely to be sufficient water available. In other words, when people have more water to wash in, they are able to wash properly, which significantly improves the survival chances of their children. Effects in individual studies of hygiene also appeared related to water supply access. For example, in Côte d’Ivoire [33], hygiene education was provided alongside village water pumps which gave 76 cubic metres per day for a community of 400 people, equivalent to 190 litres per capita per day. The study with the smallest effect on diarrhoea mortality was conducted among communities where some households had access to running water for only two hours each week [65].

Latrine promotion to whole communities was most consistently associated with the reductions in diarrhoea mortality in childhood, although we note the small number of intervention effects available (n=2). Thus, when sanitation is available and used by the majority of people in a community, it lessens children’s interactions with faeces in the public realm, reducing infection transmission and the mortality risk. In contrast, the effect on all-cause or diarrhoea mortality of household water treatment was not significant. Few studies have estimated the effects of water treatment and storage on diarrhoea mortality, and only one used a method (filtration) thought to be efficacious in removing common causes of enteric infection in low-income settings [60]. Most of the studies of HWT evaluated chlorination, which is not thought efficacious in removing common diarrhoea pathogens in low-income settings like cryptosporidium [72].

The analysis suggested WASH interventions were most effective when they were given in circumstances of high environmental risk, where most households openly defaecated or used unimproved water supply and sanitation amenities, and the baseline mortality rate was consequently higher. WASH interventions were also more effective in the summer rainy season, when environmental contamination is thought greater in warm country contexts. Diarrhoea mortality in South Asia and sub-Saharan Africa has been shown as largely associated with E. Coli infection in infants and cryptosporidium in children [73], both of which are expected to be more prevalent in warmer conditions. Shorter trials, which are usually conducted in the peak diarrhoea season when the intervention is most efficacious, also tended to have significantly larger effects on all-cause mortality. Because the season of data collection was already accounted for in meta-regression analysis, this suggested there may be other reasons for studies with longer follow-ups to have smaller effects, such as maintenance faults in the WASH technology and/or reduced adherence over time.

Meta-regression analysis suggested approximately three-quarters of deaths in childhood could be averted when WASH interventions are provided to immunocompromised groups during the peak diarrhoea season, against counterfactuals living in very poor communities with unimproved sanitation services. We found no evidence of publication bias due to small-study effects in trials of WASH interventions, presumably because mortality was not defined as an outcome in these studies.

### What the study adds to existing research

These results support predictions from theory. One would expect a stronger relationship between improved WASH access and diarrhoea mortality, than all-cause mortality, as we have found. Inadequate WASH may cause death in young children through other routes such as respiratory infection and under-nutrition, but diarrhoea is thought the biggest single cause [4]. The findings are therefore consistent with the main causes of mortality in childhood: domestic hygiene is the common factor which can block transmission of faeco-oral and respiratory infections [74]; community-wide sanitation breaks transmission of diarrhoea from open defaecation in the public and domestic domains [75]. These effects would tend to be greater over a counterfactual where existing water supply and sanitation services are not available or unimproved, so that community members are not able to practice hand washing and are openly defaecating, or using facilities that are either shared between two or more households or ones that do not adequately separate excreta from the environment.

Therefore, the significantly greater impacts of WASH interventions in contexts where environmental contamination and the baseline mortality are high, and the greater significance of findings when WASH interventions were provided together, are consistent with the WASH ladders concept: where the WASH improvement is from a lower base, or enhances access to water, sanitation and hygiene together, one would expect bigger effects on health. The review’s findings of null effects on all-cause mortality for study participants aged over 5 years is also consistent with the maturation of immunity systems with age, causing older children and adults to be less susceptible to infectious disease mortality than under-5s [76]. This finding is in contrast to reviews of diarrhoea morbidity that have found significant effects for those aged over 5 years too [77]. The F-diagram includes six intermediate transmission vectors (fluids, fields, flies, fingers, food and fomites), of which only the fluids route is addressed through water quality [78]. While we did not find significant effects, on all-cause or diarrhoea mortality, of water treatment interventions, which act on water quality, where drinking water of quality is used to prepare food it may help address food-borne transmission, thought particularly important for weaning children [79].

Non-randomised studies at ‘high risk of bias’ can produce inflated effects, as we found here, because *p*-hacking would tend to increase effect size magnitudes. However, we estimated the opposite effect for RCTs – that ‘high risk of bias’ is associated with smaller effects on mortality – a finding which is consistent with site selection bias [80,81]. In other words, trials that are more carefully conducted and reported are of interventions that also tend to be designed and implemented appropriately to the local context, and therefore adhered to, hence being more effective. An example is when interventions promote hand washing (e.g., education, social marketing, soap provision) in contexts where the quantity of water available to households is sufficient to practice domestic cleanliness; or, if it is thought not to be, improvements in water supply access or reliability are made too.

### Findings in relation to other systematic reviews

The evidence presented here, that water supply, hygiene improvements and community-wide sanitation save children’s lives in L&MICs, is consistent with findings from an early review [82], but in several respects is quite different from later reviews. These have not tended to find significant effects on diarrhoea morbidity of interventions which aim to improve access to water in quantity for household use. The most recent review by the WHO suggests that clean drinking water provided at the point-of-use, particularly by filtration, reduces reported diarrhoeal illness by around one-half [11]. Reviews have found that HWT appears to be more effective when a protective container is also provided [83], as it may be for example in household filtration devices when drinking water is accessed through a straw or tap. Reviews have also found smaller or null effects for household water treatment technologies like chlorination, when studies were double-blinded [13,23,83], or when methods were used to correct for lack of blinding [84,85]. Hand hygiene interventions have been found to have varying effects on diarrhoeal illness [74,86,87] and a review is underway to update the evidence on respiratory infection [88]. The difference between our findings for mortality and the reviews of morbidity might arise because of the contexts in which the studies have been conducted and specifically the availability of treatment. However, many of the papers and contexts included in this review are also represented in the reviews of morbidity.

A few other published reviews have provided estimates of mortality reduction due to factors associated with WASH provision. Morris et al. [89] reviewed evidence on cause-specific mortality among under-5s, estimating 22 percent of deaths were due to diarrhoea and 20 percent to pneumonia. Benova et al. [90] estimated significant reductions in maternal mortality due to improvements in water and sanitation, which appeared most closely related to water supply access (OR=0.42, 95%CI=0.29, 0.83, I-squared=0%, 2 estimates).

### Limitations of the study

The reporting of children’s deaths through interviews with mothers is susceptible to some biases and omissions, which have been investigated and documented in the literature [91,92]. Omissions are relatively common in the reporting of deaths occurring 10 to 15 years before a survey takes place, but there is no evidence of underreporting of deaths for more recent time periods. As for biases, there is no evidence that mothers from a variety of countries tend to underreport deaths occurring soon after birth or deaths of girls. Given the relatively shorter recall period employed in the studies considered in our review, we believe underreporting of deaths is unlikely. It is also not obvious why underreporting of deaths should differ between treated and untreated groups.

Hence, regarding the quality of the evidence collected here, reported mortality is not thought to be a biased measure *per se.* All-cause mortality data can also be triangulated with corresponding data from other sources, such as vital registration, and even the possible effect of other diseases, such as respiratory infections [93]. Cause-specific death rates are thought less reliable [16], dependent as they are on a verbal autopsy interview with the bereaved family of the patient, who may be too distraught to give an unbiased, let alone a coherent account of the patient’s last days. But, like all-cause mortality, verbal autopsy can be triangulated with, or done by, a physician, which we incorporated in the risk-of-bias assessment. Vital registration and verbal autopsy estimates are also used in GBD calculations.

A potentially more serious source of bias is differential attrition. During survey interviews deaths will not be reported for mothers who migrated or died. To the extent that WASH interventions affect migration and adult mortality rates, child mortality rates might be downwards biased in intervention areas. In other words, a potential source of bias affecting the crude death rate calculations used in this study is that they are right-censored: that is, where data are collected contemporaneously among participants regardless of age, children born into the study or who migrate out and younger children will have completed shorter durations than older children; the data on pre-and neo-natal mortality may also be right censored by maternal deaths in pregnancy or labour. This causes downwards bias in the estimate of mortality in any single trial arm, although the bias may be less problematic in randomised trials with contemporaneous data collection across arms. A final source of bias in mortality estimates is where severely ill children were given ORS or encouraged to attend health clinic [37,40,56–58,60,64]. Hence, for all of these reasons the results should be interpreted as providing lower-bound estimates of the impacts of WASH on mortality in childhood.

The evidence synthesis combined a variety of WASH technologies, promotional interventions, and counterfactuals. We attempted to address potential sources of inconsistency through the stratified meta-analysis by WASH technologies which incorporated information about the counterfactual scenarios. However, inconsistency of the interventions is an important potential limitation of meta-analyses of general WASH improvements. For example, we included many promotional approaches, including hygiene social marketing [34], community-led total sanitation (CLTS) [32] and latrine promotion with subsidies [53], the decentralisation of water services to local government [67] and the privatisation of local water supply and sanitation provision [50]. This inconsistency may be addressed through systematic analysis of adherence to measure actual exposures to improved WASH technologies [94], and as more studies and participant flows become available for syntheses of particular interventions.

## What the findings imply for policy and research

In 2016, the UN proclaimed 2018-2028 the International Decade for Action on Water for Sustainable Development (https://www.unwater.org/new-decade-water/). Our results provide evidentiary support for greater attention to ensuring populations can access and use improved water supplies for domestic hygiene and sanitation. We present evidence that suggests these simple interventions may significantly improve survival in early childhood from infection. Even though the review was restricted to endemic disease circumstances, the findings may also be relevant for epidemic disease control including coronavirus 2019 [95]. It is well-known that water supplies and sanitation are pro-poor and gender-inclusive interventions due to the time-savings and safety they may enable [96,97]. Our results suggest significant contributions could be made to reducing the global disease burden for diarrhoea and respiratory infections from improvements in water supplies, hygiene and sanitation where access is particularly inadequate, especially in sub-Saharan Africa and parts of South Asia.

Transparent study reporting is crucial for accountability and learning by enabling effects for relevant outcomes to be measured. A common source of bias in WASH trials is caused by differential losses to follow-up out of the study (attrition). How much attrition there is, and the reasons for it – for example, participant deaths – should be known. Reporting standards are well-known in health research due to the work of the Consolidated Standards of Reporting Trials (CONSORT) Group [98,99], and standards are available in development economics too [100]. Many authors and journals do now report this information, but there are lags in practices across the research communities producing WASH trials. According to a recent survey, participant flows have been reported in around half of studies in environmental health, but they are not typically provided in studies in development economics [10].

Water is an important enabling factor for practising hand and food hygiene and some forms of sanitation (e.g., flush toilets), but articles do not typically report data on distance to the water source, or water consumption (litres per capita per day) and how it is used (e.g., whether consumed or used in bathing). This information is crucial for understanding mechanisms and therefore the generalisability of the findings. Three studies provided information on distance to the water supply [8,32,55], one of which also reported water consumption [32]. In addition, it was not always clear exactly which interventions were provided to participants, not just the nature of the water supply improvement but whether hand or food hygiene were promoted. Therefore, a final recommendation is for more transparent reporting about the conditions being compared, including clearer information about the WASH technology itself that is being promoted and the counterfactual scenario. For example, if hygiene messaging is part of the intervention, it should be clearly indicated in the title or abstract.

## 5. Conclusions and suggested research directions

We found large and consistent effects of water supply improvements on all-cause mortality in childhood, and of community-wide sanitation improvements on diarrhoea mortality. The contribution of this synthesis – to use participant flow data to provide estimates of changes in child mortality associated with WASH interventions – has been enabled by studies that use agreed standards of reporting such as CONSORT. There is potentially a large number of estimates of mortality in childhood from studies which do not use these methods of reporting, as a recent meta-analysis of household water treatment has indicated [101]. Going forward, the challenge will be for an author collaborative to provide sufficient incentives to obtain unpublished participant flow data, to ensure that future systematic reviews and meta-analyses are representative of the complete data available on mortality in WASH intervention studies. There is also a need for more rigorous studies of water supply improvements. Although prospective evaluations of water supply interventions are being done (e.g., [102]), we are only aware of one published randomised field trial of a water supply improvement in Ghana [103] and one study that randomised encouragement of subsidies for household connections in Morocco [104]. If services are allocated by administrative area or according to a threshold rule (e.g., the share of community members currently covered by a service), it may be possible to use a discontinuity design, an approach that has been shown to generate the same effect estimates as RCTs, whether applied prospectively or retrospectively [105]. We are hopeful that the evidence presented in this review, and the evidence we are calling for, will prove useful for those taking decisions about what WASH improvements are needed in the second half of the International Decade for Action on Water.

## Data Availability

All relevant data are within the manuscript and its Supporting Information files.

## Acknowledgements

Helpful comments were given by participants at UNC Water and Health 2021, the What Works Global Summit 2021, the LSHTM-IFS WASH Economics Conference in September 2022, the University of Salford, Britta Augsburg, Paul Hunter and our colleagues at the Environmental Health Group at LSHTM.

## Supporting information

S1 Annex tables

S2 PRISMA 2020 checklist S3 Annex figures

S4 Dataset

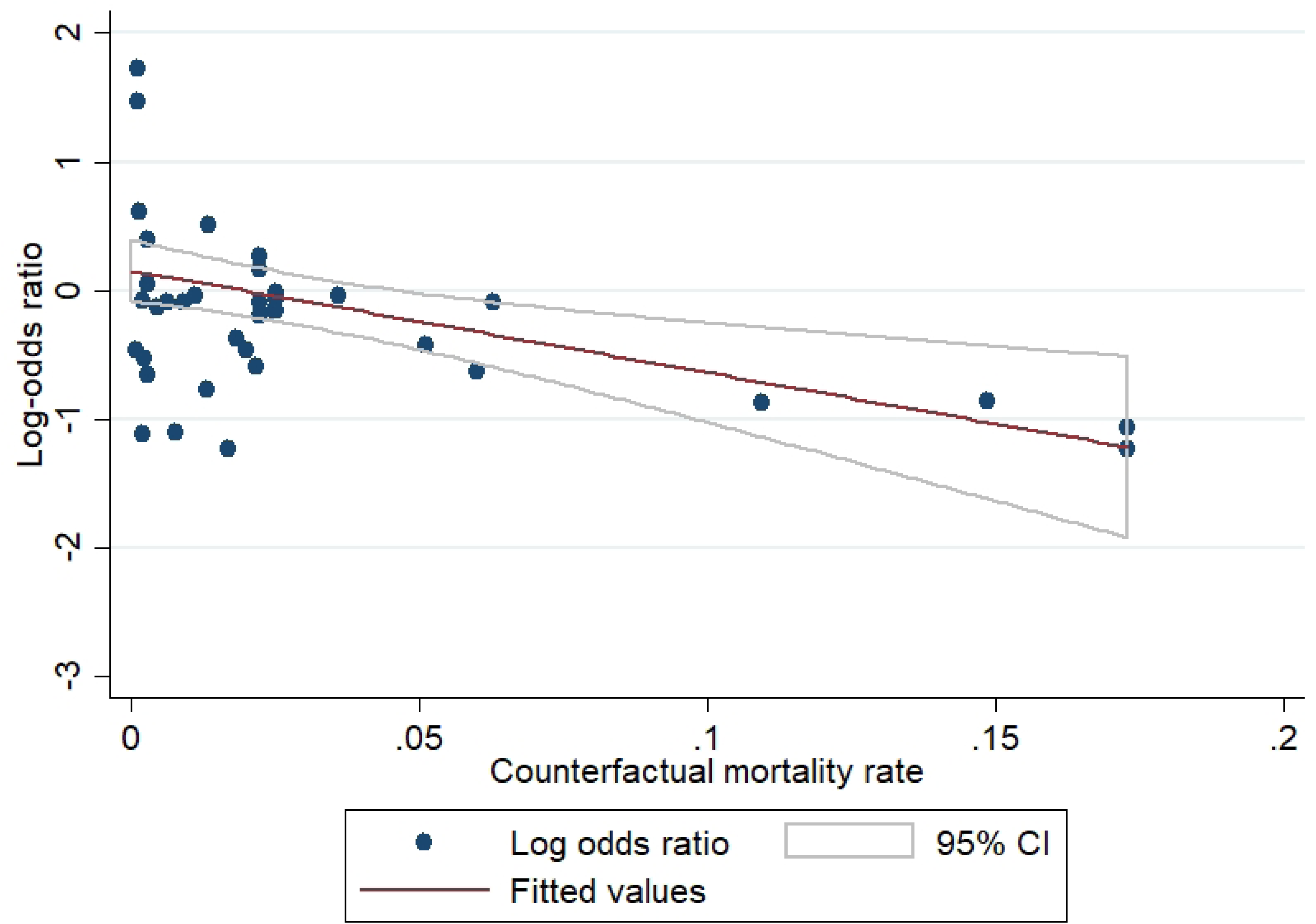

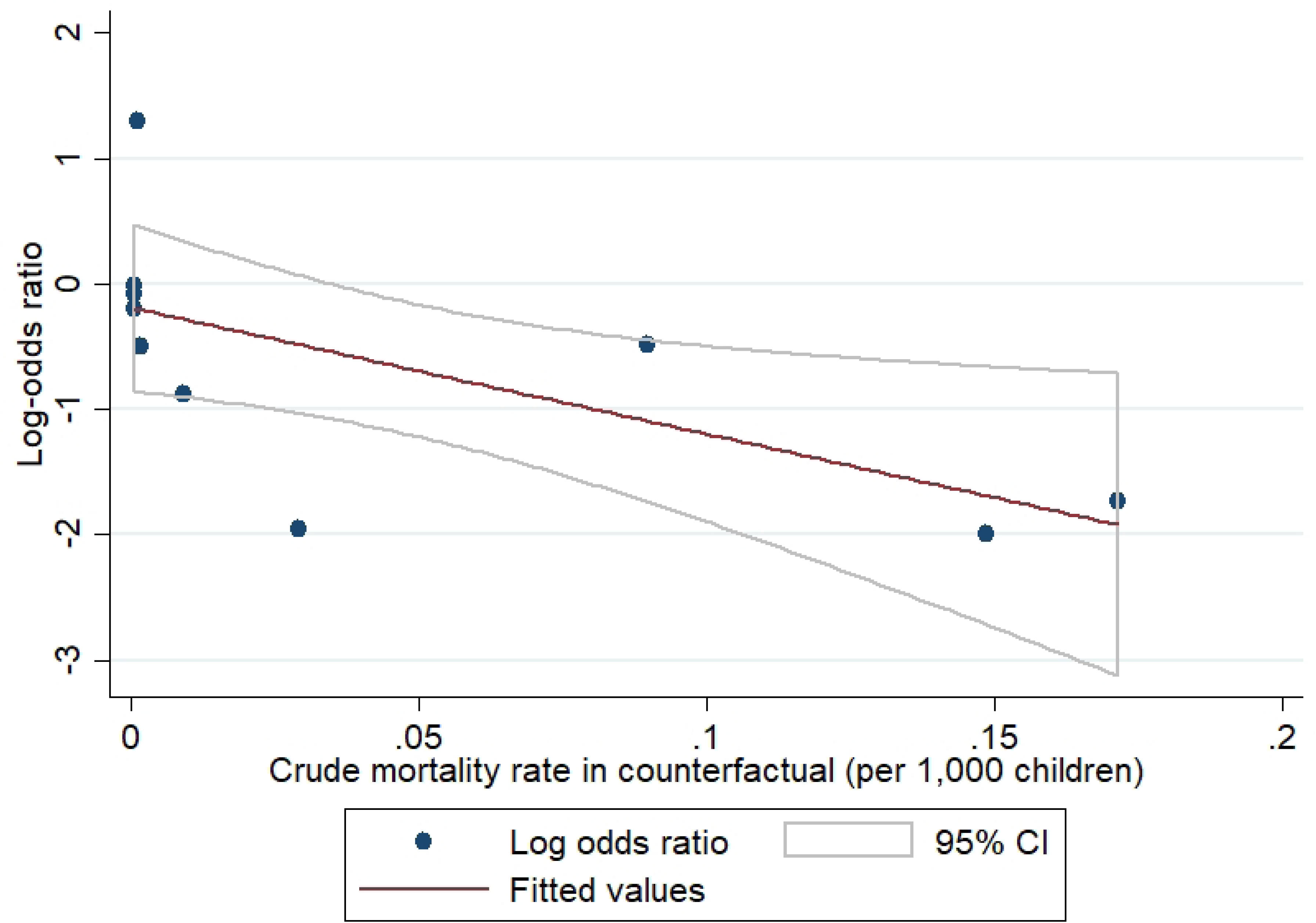

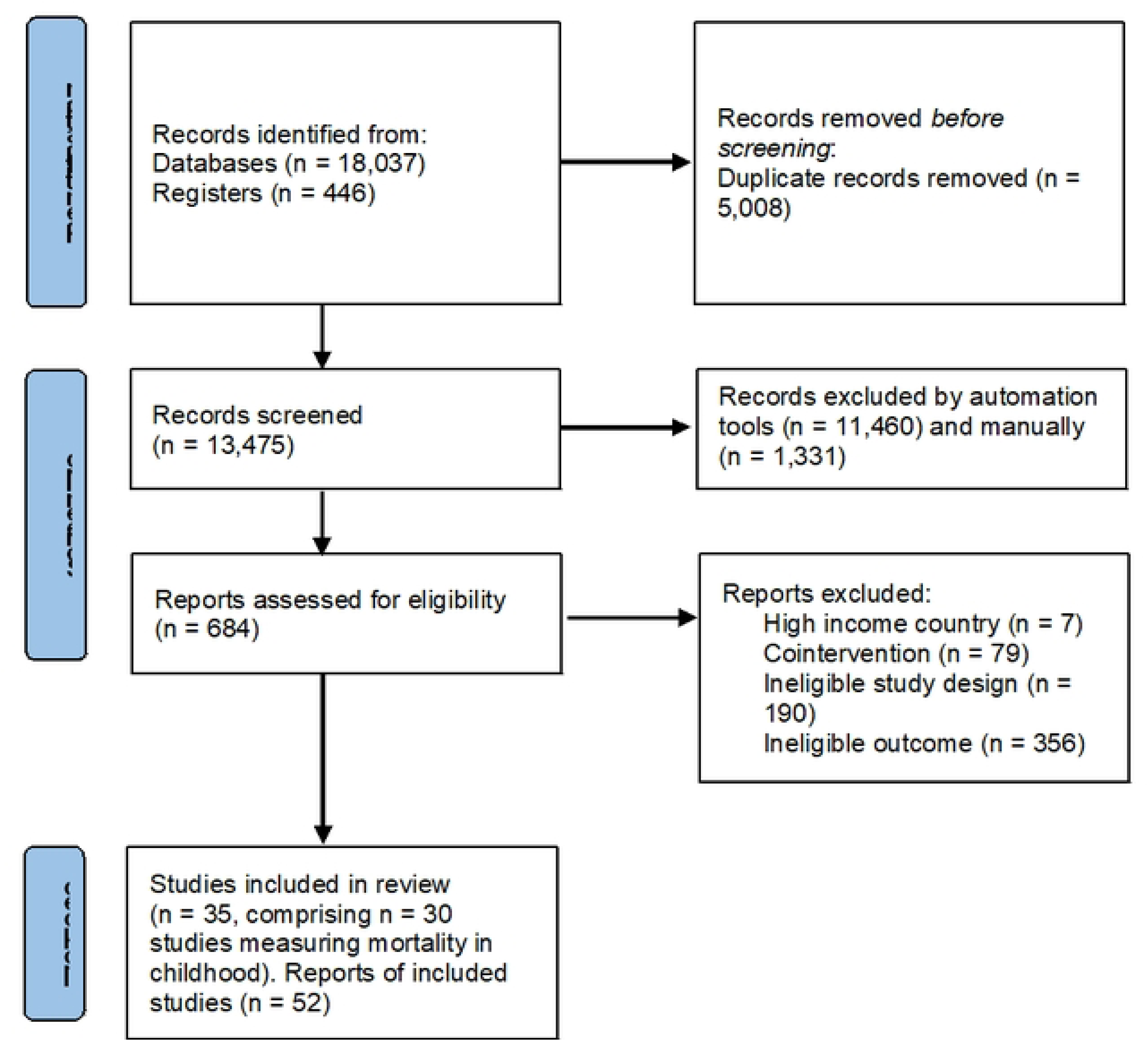

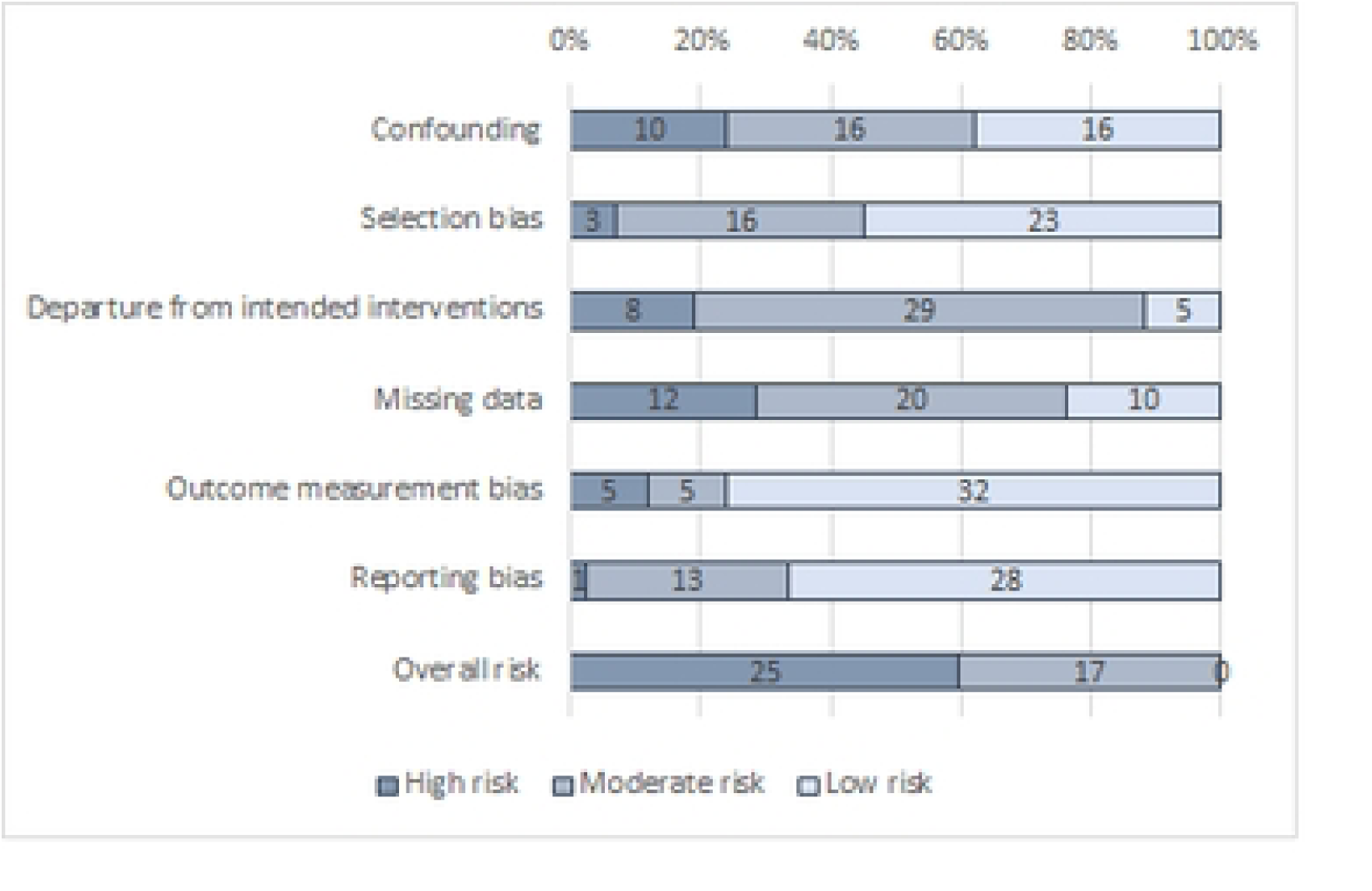

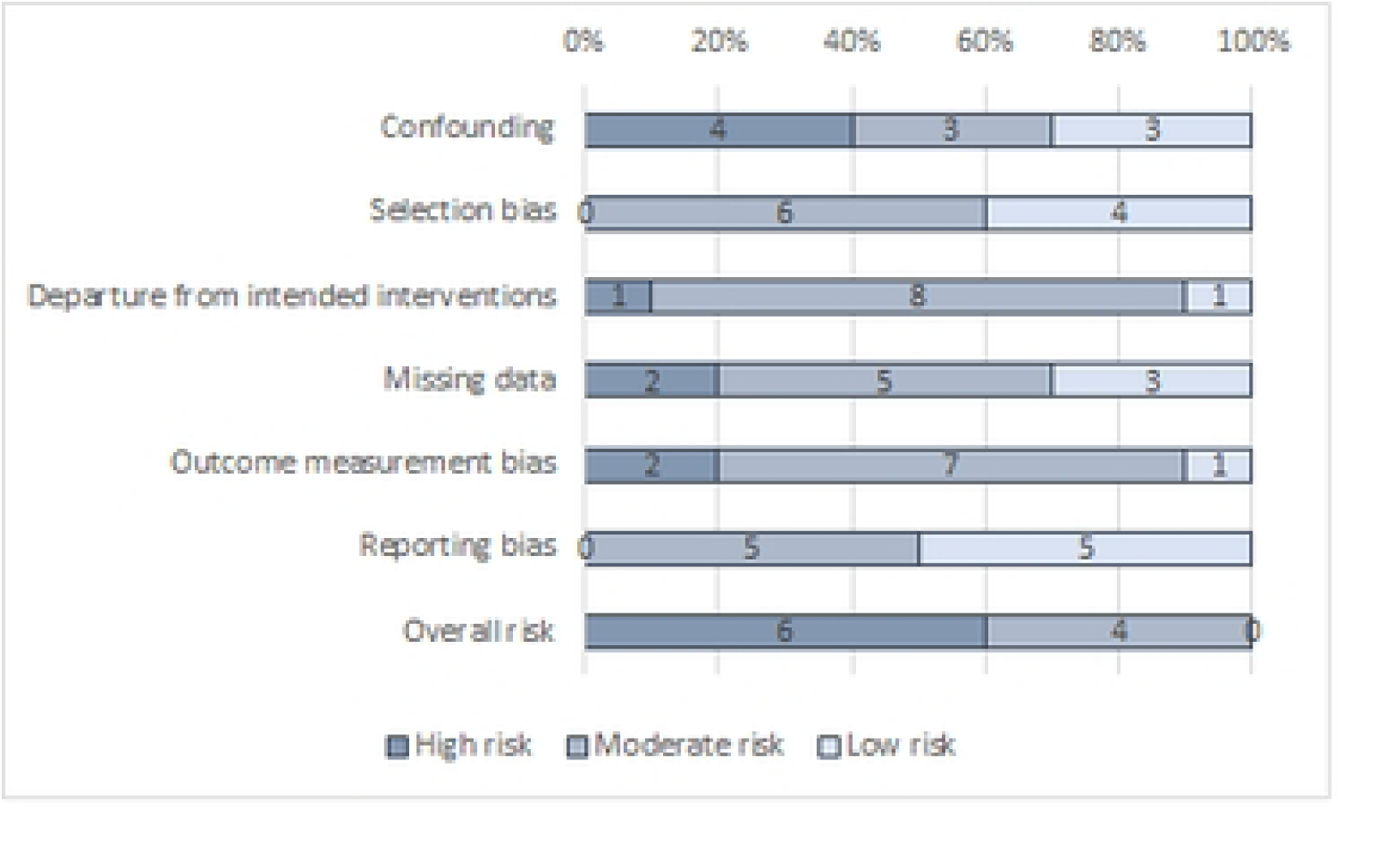

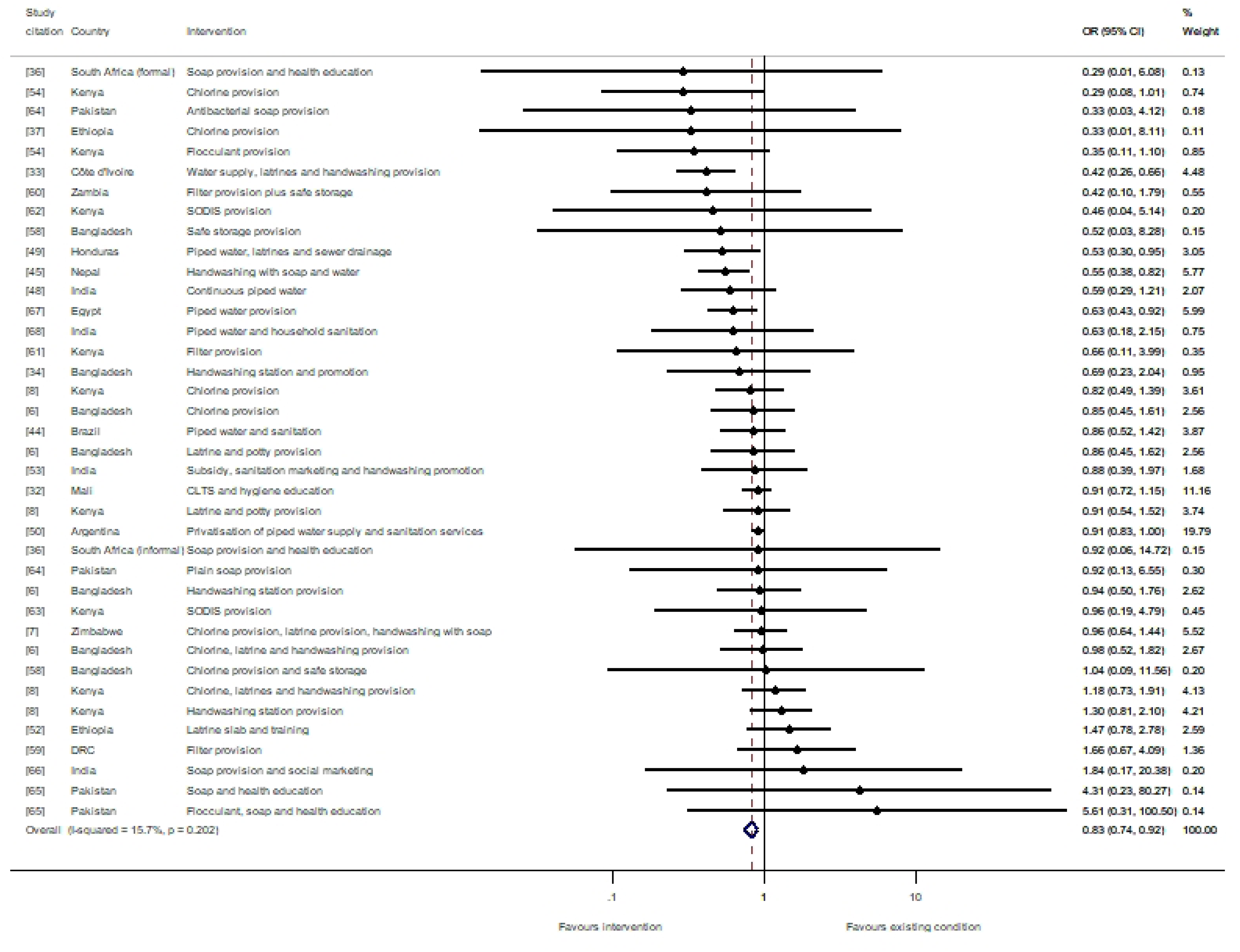

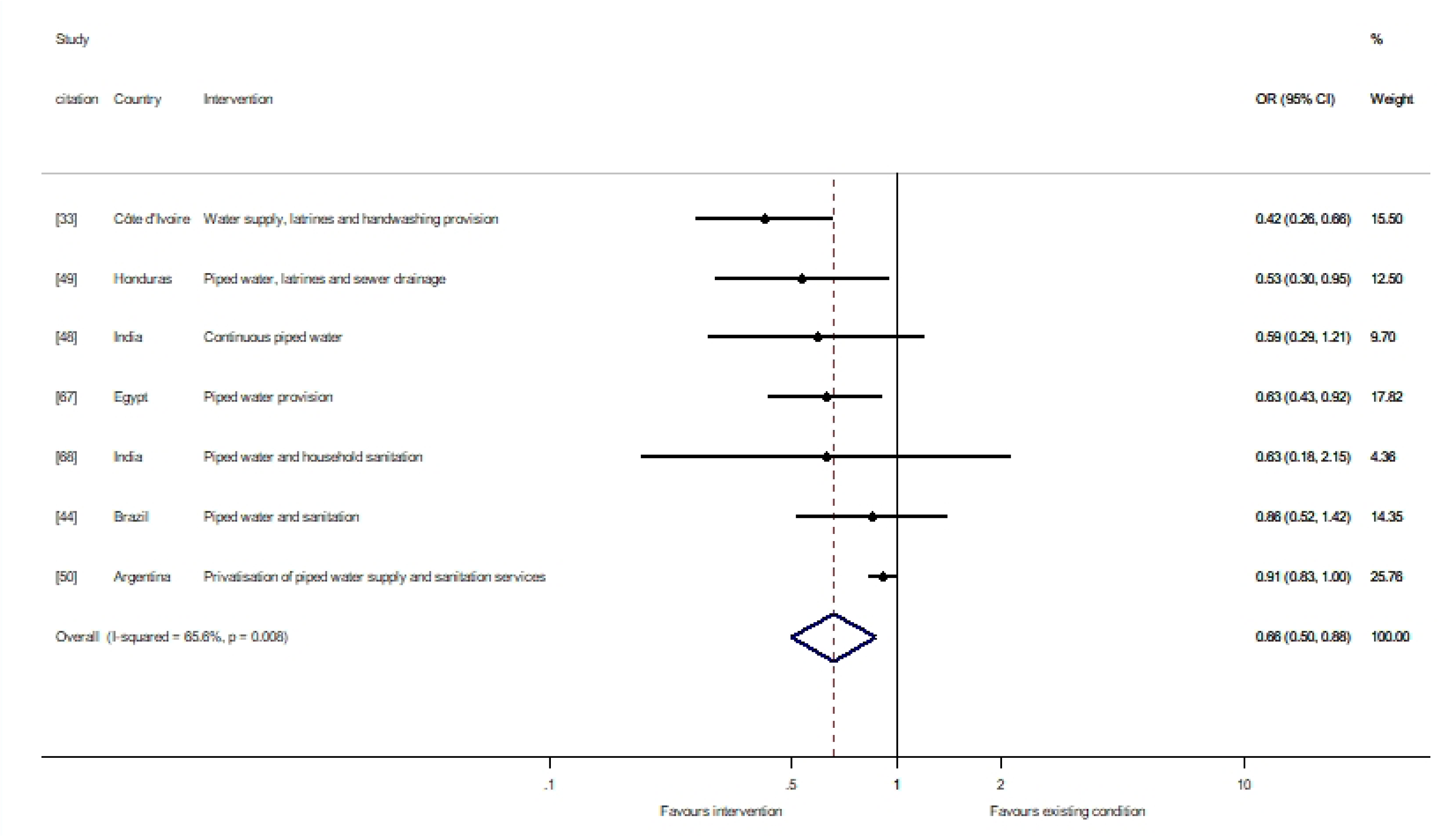

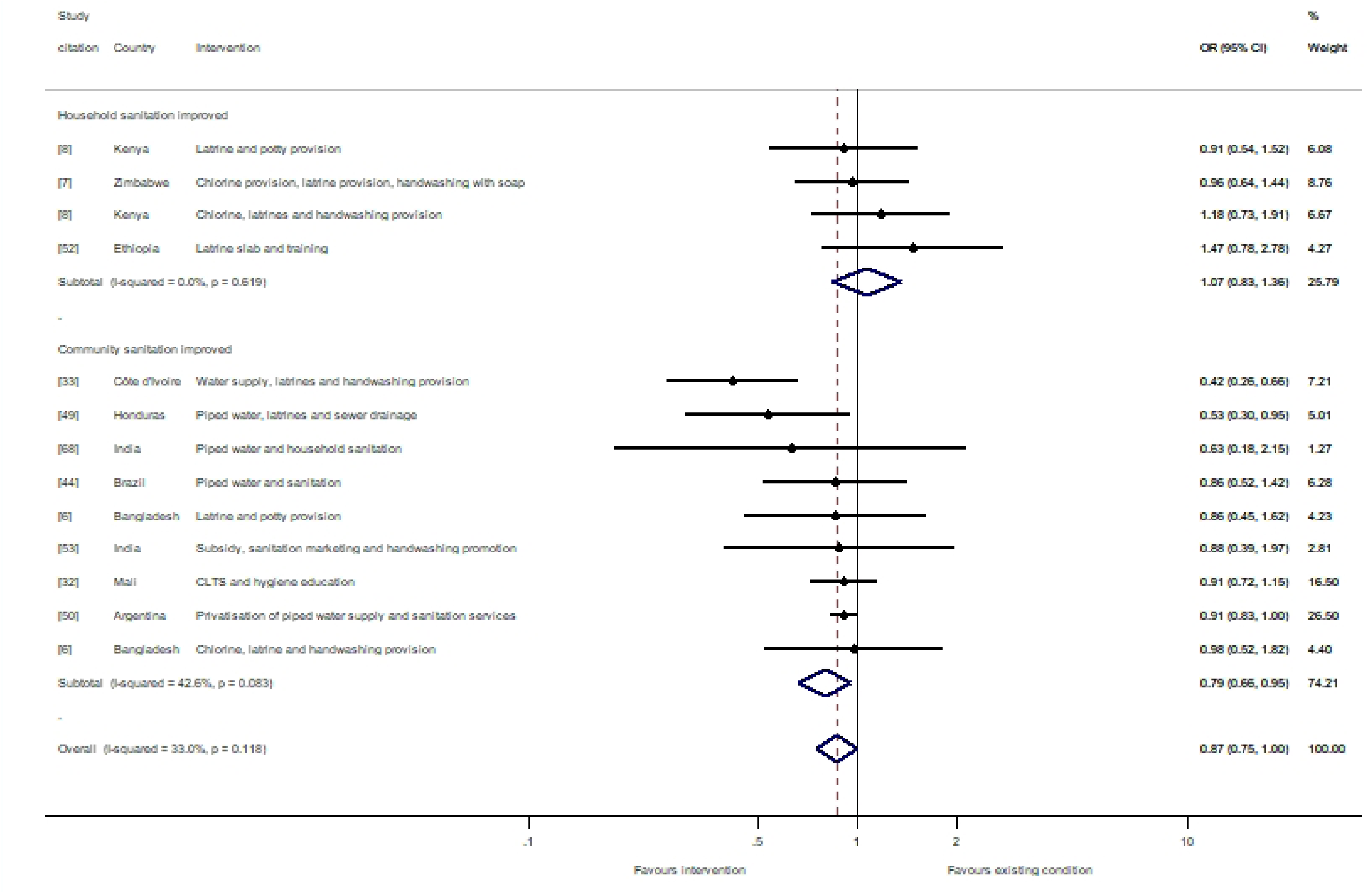

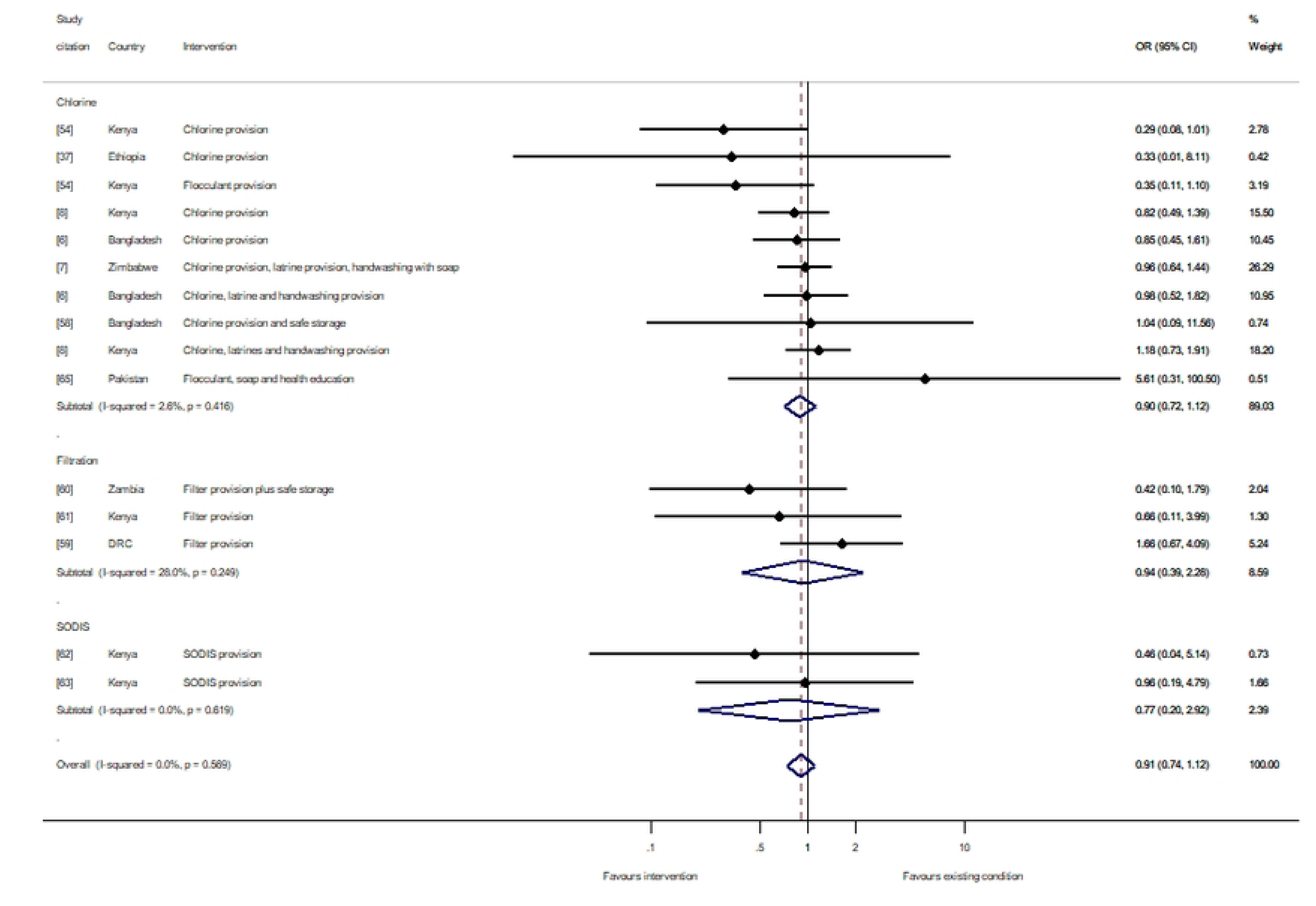

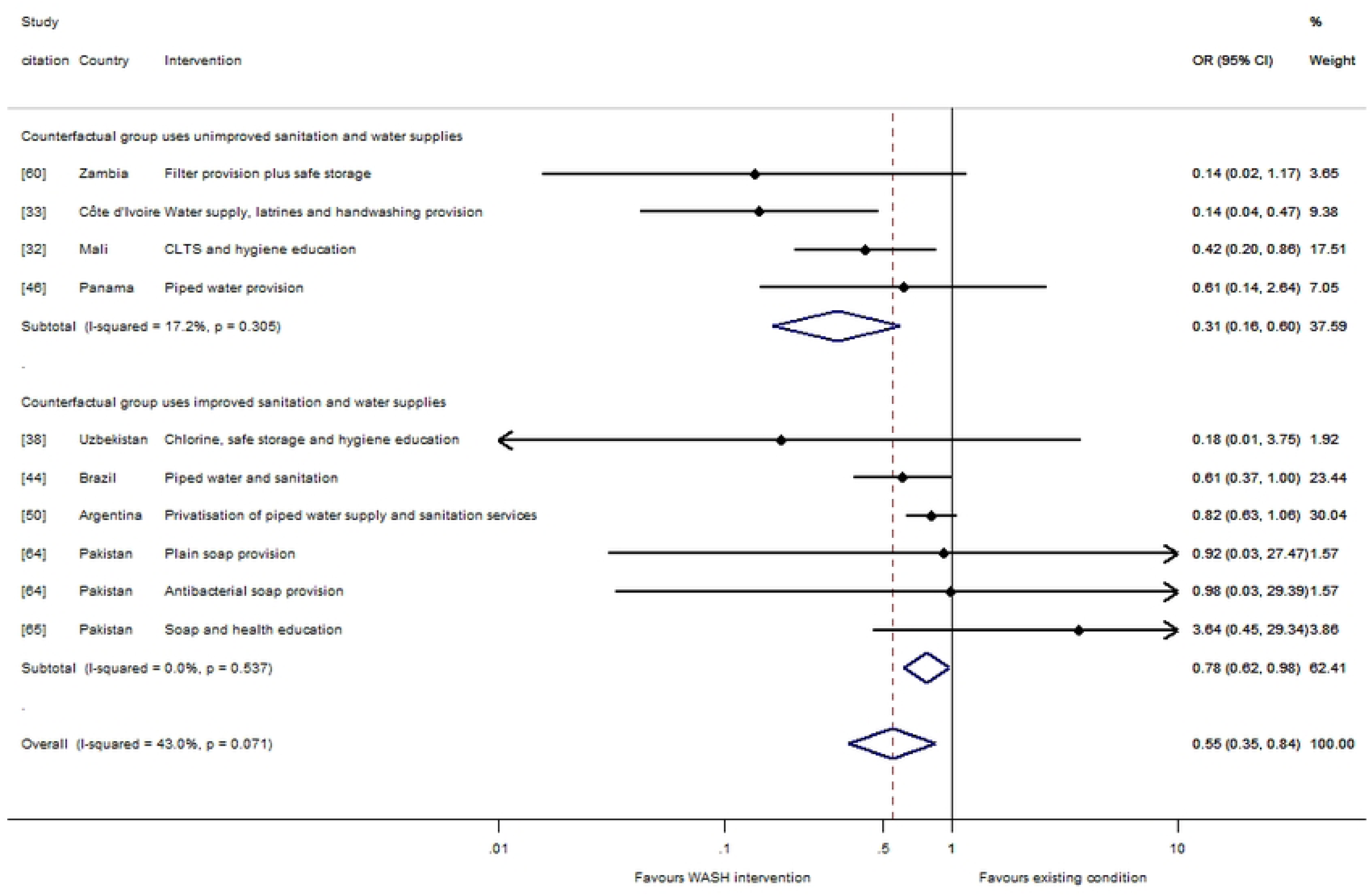

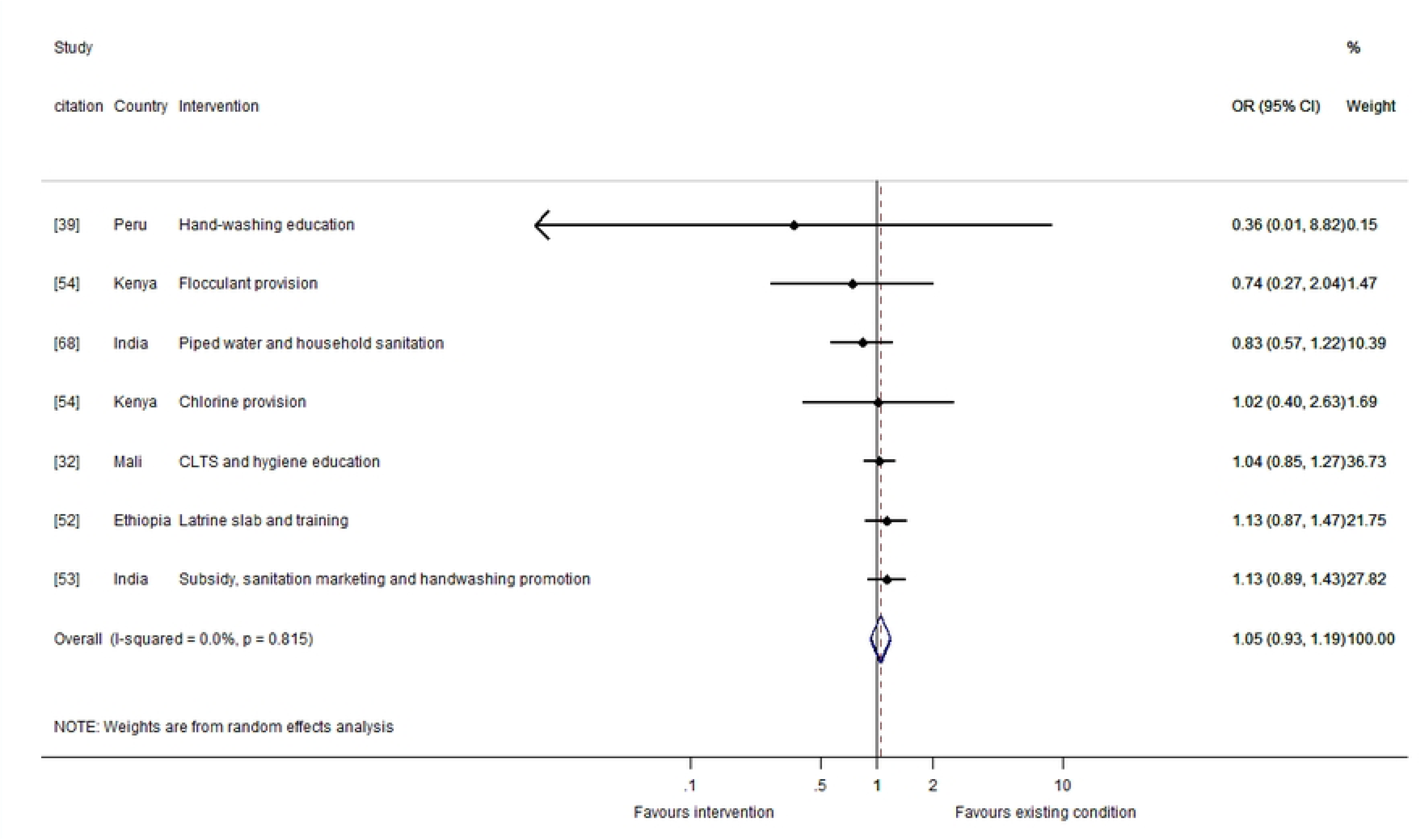

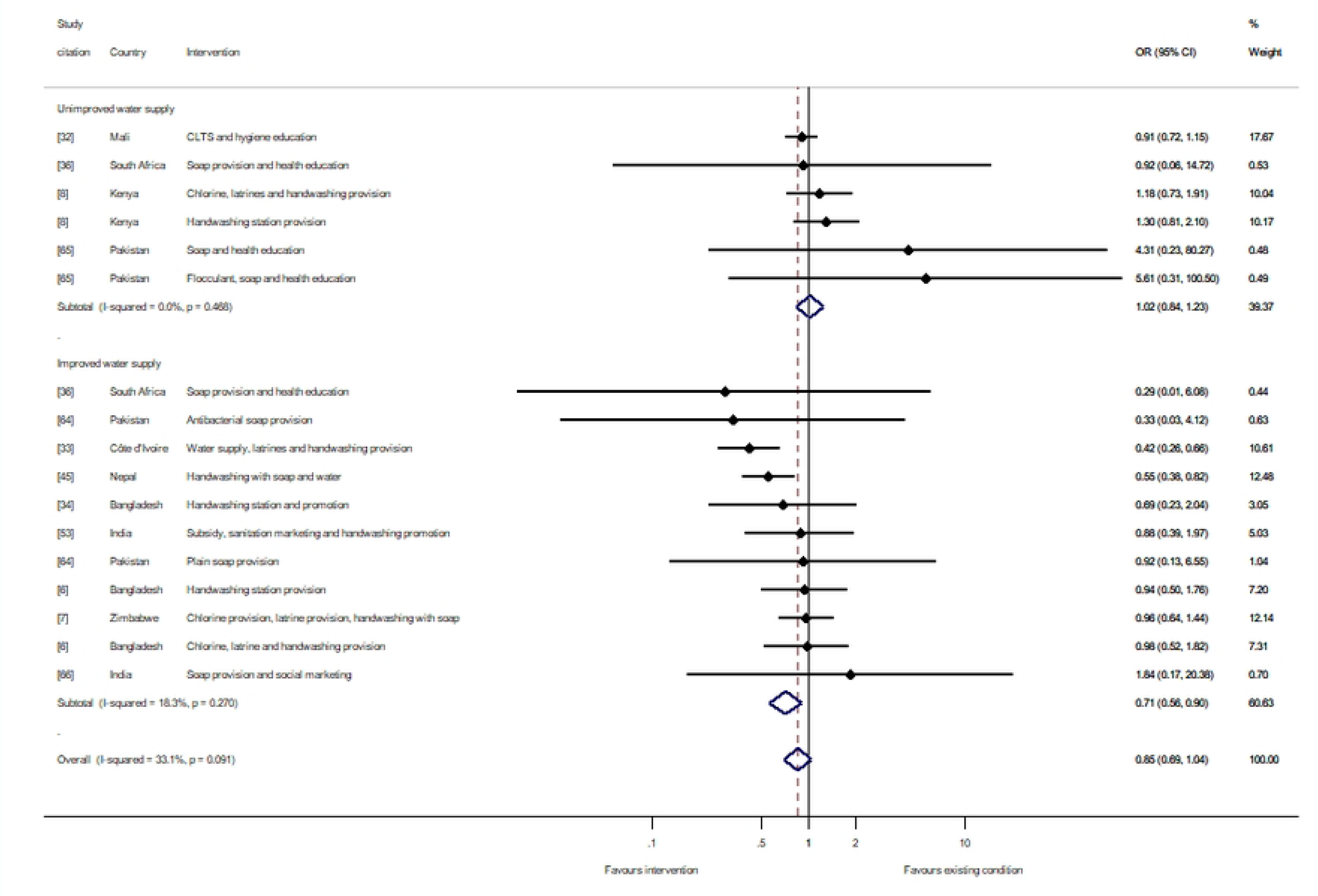

## References

1. Naghavi M, Abajobir AA, Abbafati C, Abbas KM, Abd-Allah F, Abera SF, et al. Global, regional, and national age-sex specific mortality for 264 causes of death, 1980–2016: a systematic analysis for the Global Burden of Disease Study 2016. The lancet. 2017;390(10100):1151–210.

2. Troeger C, Blacker BF, Khalil IA, Rao PC, Cao S, Zimsen SR, et al. Estimates of the global, regional, and national morbidity, mortality, and aetiologies of diarrhoea in 195 countries: a systematic analysis for the Global Burden of Disease Study 2016. Lancet Infect Dis. 2018;18(11):1211–28.

3. Walker CLF, Rudan I, Liu L, Nair H, Theodoratou E, Bhutta ZA, et al. Global burden of childhood pneumonia and diarrhoea. Lancet Lond Engl. 2013 Apr 20;381(9875):1405–16.

4. Prüss-Ustün A, Wolf J, Bartram J, Clasen T, Cumming O, Freeman MC, et al. Burden of disease from inadequate water, sanitation and hygiene for selected adverse health outcomes: An updated analysis with a focus on low-and middle-income countries. Int J Hyg Environ Health. 2019 Jun;222(5):765–77.

5. Victora CG, Smith PG, Vaughan JP, Nobre LC, Lombard C, Teixeira AMB, et al. Water supply, sanitation and housing in relation to the risk of infant mortality from diarrhoea. Int J Epidemiol. 1988;17(3):651–4.

6. Luby SP, Rahman M, Arnold BF, Unicomb L, Ashraf S, Winch PJ, et al. Effects of water quality, sanitation, handwashing, and nutritional interventions on diarrhoea and child growth in rural Bangladesh: a cluster randomised controlled trial. Lancet Glob Health. 2018;6(3):e302–15.

7. Humphrey JH, Mbuya MN, Ntozini R, Moulton LH, Stoltzfus RJ, Tavengwa NV, et al. Independent and combined effects of improved water, sanitation, and hygiene, and improved complementary feeding, on child stunting and anaemia in rural Zimbabwe: a cluster-randomised trial. Lancet Glob Health. 2019;7(1):e132–47.

8. Null C, Stewart CP, Pickering AJ, Dentz HN, Arnold BF, Arnold CD, et al. Effects of water quality, sanitation, handwashing, and nutritional interventions on diarrhoea and child growth in rural Kenya: a cluster-randomised controlled trial. Lancet Glob Health. 2018 Mar 1;6(3):e316–29.

9. Guyatt G, Oxman AD, Akl EA, Kunz R, Vist G, Brozek J, et al. GRADE guidelines: 1. Introduction—GRADE evidence profiles and summary of findings tables. J Clin Epidemiol. 2011;64(4):383–94.

10. Chirgwin H, Cairncross S, Zehra D, Sharma Waddington H. Interventions promoting uptake of water, sanitation and hygiene (WASH) technologies in low-and middle-income countries: An evidence and gap map of effectiveness studies. Campbell Syst Rev. 2021;17(4):e1194.

11. Wolf J, Hubbard S, Brauer M, Ambelu A, Arnold BF, Bain R, et al. Effectiveness of interventions to improve drinking water, sanitation, and handwashing with soap on risk of diarrhoeal disease in children in low-income and middle-income settings: a systematic review and meta-analysis. The Lancet. 2022;400(10345):48–59.

12. Fewtrell L, Colford JMJ. Water, Sanitation and Hygiene : Interventions and Diarrhoea [Internet]. Washington, DC: World Bank; 2004 Jul [cited 2022 Sep 15]. Available from: https://openknowledge.worldbank.org/handle/10986/13742

13. Schmidt WP, Cairncross S. Household water treatment in poor populations: is there enough evidence for scaling up now? Environ Sci Technol. 2009;43(4):986–92.

14. Zwane AP, Zinman J, Van Dusen E, Pariente W, Null C, Miguel E, et al. Being surveyed can change later behavior and related parameter estimates. Proc Natl Acad Sci. 2011;108(5):1821–6.

15. Wood L, Egger M, Gluud LL, Schulz KF, Jüni P, Altman DG, et al. Empirical evidence of bias in treatment effect estimates in controlled trials with different interventions and outcomes: meta-epidemiological study. Bmj. 2008;336(7644):601–5.

16. Savović J, Jones HE, Altman DG, Harris RJ, Jüni P, Pildal J, et al. Influence of reported study design characteristics on intervention effect estimates from randomised controlled trials: combined analysis of meta-epidemiological studies. Health Technol Assess. 2012;16(35):1–82.

17. Clasen T. Comments from Dr. Thomas Clasen on a draft of the GiveWell Water Quality Report, November 15, 2013 [Internet]. GiveWell; 2013 [cited 2019 Jan 1]. Available from: https://files.givewell.org/files/DWDA%202009/Interventions/Water/Water%20Purification%20Assessment/eBeer-clasen-comments.pdf

18. Schmidt WP, Arnold BF, Boisson S, Genser B, Luby SP, Barreto ML, et al. Epidemiological methods in diarrhoea studies—an update. Int J Epidemiol. 2011 Dec;40(6):1678–92.

19. Briscoe J, Feachem RG, Rahaman MM. Evaluating health impact: water supply, sanitation, and hygiene education. IDRC, Ottawa, ON, CA; 1986.

20. Imo State Evaluation Team. Evaluating water and sanitation projects: lessons from Imo State, Nigeria. Health Policy Plan. 1989;40–9.

21. Deaton A, Cartwright N. Understanding and misunderstanding randomized controlled trials. Soc Sci Med. 2018;210:2–21.

22. Higgins JP, Thomas J, Chandler J, Cumpston M, Li T, Page M, et al. Cochrane handbook for systematic reviews of interventions version 6.2 (updated February 2021) [Internet]. Cochrane; 2021 [cited 2021 May 31]. Available from: https://training.cochrane.org/handbook/current

23. Waddington H, Snilstveit B, White H, Fewtrell L. Water, sanitation and hygiene interventions to combat childhood diarrhoea in developing countries. New Delhi Int Initiat Impact Eval. 2009;

24. Sharma Waddington H, Cairncross S. PROTOCOL: Water, sanitation and hygiene for reducing childhood mortality in low-and middle-income countries. Campbell Syst Rev. 2021;17(1):e1135.

25. Thomas J, Brunton J, Graziosi S. EPPI-Reviewer 4.0: software for research synthesis. EPPI-Centre Software. London: Social Science Research Unit. Inst Educ Univ Lond. 2010;20(18):8–11.

26. Bongartz P. CLTS Update February 2012 [Internet]. Institute of Development Studies; 2012 [cited 2021 Nov 1]. Available from: https://archive.ids.ac.uk/clts/sites/communityledtotalsanitation.org/files/CLTS_Update_Feb_2012.pdf

27. Higgins JP, Sterne JA, Savovic J, Page MJ, Hróbjartsson A, Boutron I, et al. A revised tool for assessing risk of bias in randomized trials. Cochrane Database Syst Rev. 2016;10(Suppl 1):29–31.

28. Eldridge S, Campbell M, Campbell M, Drahota-Towns A, Giraudeau B, Higgins J, et al. Revised Cochrane risk of bias tool for randomized trials (RoB 2.0): additional considerations for cluster-randomized trials. 2016 [cited 2021 Jan 1]; Available from: https://methods.cochrane.org/bias/resources/rob-2-revised-cochrane-risk-bias-tool-randomized-trials

29. Sterne JA, Hernán MA, Reeves BC, Savović J, Berkman ND, Viswanathan M, et al. ROBINS-I: a tool for assessing risk of bias in non-randomised studies of interventions. bmj. 2016;355.

30. Waddington H, White H, Snilstveit B, Hombrados JG, Vojtkova M, Davies P, et al. How to do a good systematic review of effects in international development: a tool kit. J Dev Eff. 2012;4(3):359–87.

31. Campbell Collaboration. Campbell Collaboration Systematic Reviews: Policies and Guidelines [Internet]. The Campbell Collaboration; 2021 [cited 2022 Sep 15]. Available from: https://campbellcollaboration.org/library/campbell-collaboration-systematic-reviews-policies-and-guidelines.html

32. Pickering AJ, Djebbari H, Lopez C, Coulibaly M, Alzua ML. Effect of a community-led sanitation intervention on child diarrhoea and child growth in rural Mali: a cluster-randomised controlled trial. Lancet Glob Health. 2015 Nov 1;3(11):e701–11.

33. Messou E, Sangaré SV, Josseran R, Le Corre C, Guélain J. [Effect of hygiene measures, water sanitation and oral rehydration therapy on diarrhea in children less than five years old in the south of Ivory Coast]. Bull Soc Pathol Exot 1990. 1997;90(1):44–7.

34. Ram PK, Nasreen, S, Kamm, K, Allen, J, Kumar, S, Rahman, MA, et al. Impact of an Intensive Perinatal Handwashing Promotion Intervention on Maternal Handwashing Behavior in the Neonatal Period: Findings from a Randomized Controlled Trial in Rural Bangladesh. BioMed Res Int [Internet]. 2017 [cited 2022 Sep 15]; Available from: https://www.hindawi.com/journals/bmri/2017/6081470/

35. Efthimiou O. Practical guide to the meta-analysis of rare events. Evid Based Ment Health. 2018 May;21(2):72–6.

36. Cole EC, Hawkley M, Rubino JR, Crookston BT, McCue K, Dixon J, et al. Comprehensive family hygiene promotion in peri-urban Cape Town: Gastrointestinal and respiratory illness and skin infection reduction in children aged under 5. South Afr J Child Health. 2012;6(4):109–17.

37. Mengistie B, Berhane Y, Worku A. Household Water Chlorination Reduces Incidence of Diarrhea among Under-Five Children in Rural Ethiopia: A Cluster Randomized Controlled Trial. PLOS ONE. 2013 Oct 23;8(10):e77887.

38. Semenza JC, Roberts L, Henderson A, Bogan J, Rubin CH. Water distribution system and diarrheal disease transmission: a case study in Uzbekistan. Am J Trop Med Hyg. 1998;59(6):941–6.

39. Gyorkos TW, Maheu-Giroux M, Blouin B, Casapia M. Impact of health education on soil-transmitted helminth infections in schoolchildren of the Peruvian Amazon: a cluster-randomized controlled trial. PLoS Negl Trop Dis. 2013;7(9):e2397.

40. Luby SP, Agboatwalla M, Painter J, Altaf A, Billhimer W, Keswick B, et al. Combining drinking water treatment and hand washing for diarrhoea prevention, a cluster randomised controlled trial. Trop Med Int Health. 2006;11(4):479–89.

41. Lipsitch M, Tchetgen ET, Cohen T. Negative controls: a tool for detecting confounding and bias in observational studies. Epidemiol Camb Mass. 2010;21(3):383.

42. Arnold BF, Ercumen A. Negative Control Outcomes: A Tool to Detect Bias in Randomized Trials. JAMA. 2016 Dec 27;316(24):2597–8.

43. Kremer M, Leino J, Miguel E, Zwane AP. Spring Cleaning: Rural Water Impacts, Valuation, and Property Rights Institutions*. Q J Econ. 2011 Feb 1;126(1):145–205.

44. Rasella D. Impacto do Programa Água para Todos (PAT) sobre a morbi-mortalidade por diarreia em crianças do Estado da Bahia, Brasil. Cad Saúde Pública. 2003;29:40–50.

45. Rhee V, Mullany LC, Khatry SK, Katz J, LeClerq SC, Darmstadt GL, et al. Maternal and Birth Attendant Hand Washing and Neonatal Mortality in Southern Nepal. Arch Pediatr Adolesc Med. 2008 Jul 7;162(7):603–8.

46. Ryder R, Reeves W, Singh N, Hall C, Kapikian A, Gomez B, et al. The childhood health effects of an improved water supply system on a remote Panamanian island. Am J Trop Med Hyg. 1985;34(5):021–924.

47. Reese H, Routray P, Torondel B, Sinharoy SS, Mishra S, Freeman MC, et al. Assessing longer-term effectiveness of a combined household-level piped water and sanitation intervention on child diarrhoea, acute respiratory infection, soil-transmitted helminth infection and nutritional status: a matched cohort study in rural Odisha, India. Int J Epidemiol. 2019 Dec 1;48(6):1757–67.

48. Ercumen A, Arnold BF, Kumpel E, Burt Z, Ray I, Nelson K, et al. Upgrading a piped water supply from intermittent to continuous delivery and association with waterborne illness: a matched cohort study in urban India. PLoS Med. 2015;12(10):e1001892.

49. Instituto Apoyo. Evaluacion de impacto y sostenibilidad de los proyectos de foncodes. Instituto Apoyo, Tegucigalpa. [Internet]. Instituto Apoyo, Tegucigalpa; 2000 [cited 2021 Feb 8]. Available from: https://fdocuments.ec/document/sexta-evaluacin-expost-del-foncodes-world-rubio-en-la-definicin-del-diseo.html?page=7

50. Galiani S, Gertler P, Schargrodsky E. Water for life: The impact of the privatization of water services on child mortality. J Polit Econ. 2005;113(1):83–120.

51. Granados C, Sánchez F. Water reforms, decentralization and child mortality in Colombia, 1990–2005. World Dev. 2014;53:68–79.

52. Gebre T, Ayele B, Zerihun M, House JI, Stoller NE, Zhou Z, et al. Latrine Promotion for Trachoma: Assessment of Mortality from a Cluster-Randomized Trial in Ethiopia. Am J Trop Med Hyg. 2011 Sep 1;85(3):518–23.

53. Clasen T, Boisson S, Routray P, Torondel B, Bell M, Cumming O, et al. Effectiveness of a rural sanitation programme on diarrhoea, soil-transmitted helminth infection, and child malnutrition in Odisha, India: a cluster-randomised trial. Lancet Glob Health. 2014 Nov 1;2(11):e645–53.

54. Crump JA, Otieno PO, Slutsker L, Keswick BH, Rosen DH, Hoekstra RM, et al. Household based treatment of drinking water with flocculant-disinfectant for preventing diarrhoea in areas with turbid source water in rural western Kenya: cluster randomised controlled trial. Bmj. 2005;331(7515):478.

55. Emerson PM, Lindsay SW, Alexander N, Bah M, Dibba SM, Faal HB, et al. Role of flies and provision of latrines in trachoma control: cluster-randomised controlled trial. The Lancet. 2004;363(9415):1093–8.

56. Lule JR, Mermin J, Ekwaru JP, Malamba S, Downing R, Ransom R, et al. Effect of home based water chlorination and safe storage on diarrhea among persons with human immonodeficiency virus in Uganda. Am J Trop Med Hyg. 2005;73(5):926–33.

57. Jain S, Sahanoon OK, Blanton E, Schmitz A, Wannemuehler KA, Hoekstra RM, et al. Sodium dichloroisocyanurate tablets for routine treatment of household drinking water in periurban Ghana: a randomized controlled trial. Am J Trop Med Hyg. 2010;82(1):16.

58. Ercumen A, Naser AM, Unicomb L, Arnold BF, Colford JMJ, Luby SP. Effects of Source versus Household Contamination of Tubewell Water on Child Diarrhea in Rural Bangladesh: A Randomized Controlled Trial. PLOS ONE. 2015 Mar 27;10(3):e0121907.

59. Boisson S, Kiyombo M, Sthreshley L, Tumba S, Makambo J, Clasen T. Field Assessment of a Novel Household-Based Water Filtration Device: A Randomised, Placebo-Controlled Trial in the Democratic Republic of Congo. PLOS ONE. 2010 Sep 10;5(9):e12613.

60. Peletz R, Simunyama M, Sarenje K, Baisley K, Filteau S, Kelly P, et al. Assessing water filtration and safe storage in households with young children of HIV-positive mothers: a randomized, controlled trial in Zambia. Am J Trop Med Hyg. 2012;101(3):555–65.

61. Morris JF, Murphy J, Fagerli K, Schneeberger C, Jaron P, Moke F, et al. A Randomized Controlled Trial to Assess the Impact of Ceramic Water Filters on Prevention of Diarrhea and Cryptosporidiosis in Infants and Young Children—Western Kenya, 2013. Am J Trop Med Hyg. 2018 May;98(5):1260–8.

62. Conroy RM, Meegan ME, Joyce T, McGuigan K, Barnes J. Solar disinfection of water reduces diarrhoeal disease: an update. Arch Dis Child. 1999;81(4):337–8.

63. du Preez M, Conroy RM, Ligondo S, Hennessy J, Elmore-Meegan M, Soita A, et al. Randomized intervention study of solar disinfection of drinking water in the prevention of dysentery in Kenyan children aged under 5 years. Environ Sci Technol. 2011;45(21):9315–23.

64. Luby SP, Agboatwalla M, Painter J, Altaf A, Billhimer WL, Hoekstra RM. Effect of Intensive Handwashing Promotion on Childhood Diarrhea in High-Risk Communities in Pakistan: A Randomized Controlled Trial. JAMA. 2004 Jun 2;291(21):2547–54.

65. Bowen A, Agboatwalla M, Luby S, Tobery T, Ayers T, Hoekstra RM. Association between intensive handwashing promotion and child development in Karachi, Pakistan: a cluster randomized controlled trial. Arch Pediatr Adolesc Med. 2012;166(11):1037–44.

66. Nicholson JA, Naeeni M, Hoptroff M, Matheson JR, Roberts AJ, Taylor D, et al. An investigation of the effects of a hand washing intervention on health outcomes and school absence using a randomised trial in Indian urban communities. Trop Med Int Health. 2014;19(3):284–92.

67. Abou-Ali H, El-Azony H, El-Laithy H, Haughton J, Khandker S. Evaluating the impact of Egyptian social fund for development programmes. J Dev Eff. 2010;2(4):521–55.

68. Reese H, Routray P, Torondel B, Sclar G, Delea MG, Sinharoy SS, et al. Design and rationale of a matched cohort study to assess the effectiveness of a combined household-level piped water and sanitation intervention in rural Odisha, India. BMJ Open. 2017;7(3):e012719.

69. Shuval HI, Tilden RL, Perry BH, Grosse RN. Effect of investments in water supply and sanitation on health status: a threshold-saturation theory. Bull World Health Organ. 1981;59(2):243.

70. White H, Menon R, Waddington H. Community-driven development: Does it build social cohesion or infrastructure? A mixed-method evidence synthesis. Working Paper 30. International Initiative for Impact Evaluation, New Delhi; 2018.

71. Otu A, Nsutebu EF, Hirst JE, Thompson K, Walker K, Yaya S. How to close the maternal and neonatal sepsis gap in sub-Saharan Africa. BMJ Glob Health. 2020 Apr 1;5(4):e002348.

72. Cumming O, Arnold BF, Ban R, Clasen T, Esteves Mills J, Freeman MC, et al. The implications of three major new trials for the effect of water, sanitation and hygiene on childhood diarrhea and stunting: a consensus statement. BMC Med. 2019 Aug 28;17(1):173.

73. Kotloff KL, Nataro JP, Blackwelder WC, Nasrin D, Farag TH, Panchalingam S, et al. Burden and aetiology of diarrhoeal disease in infants and young children in developing countries (the Global Enteric Multicenter Study, GEMS): a prospective, case-control study. The Lancet. 2013;382(9888):209–22.

74. Curtis V, Cairncross S. Effect of washing hands with soap on diarrhoea risk in the community: a systematic review. Lancet Infect Dis. 2003;3(5):275–81.

75. Cairncross S, Blumenthal U, Kolsky P, Moraes L, Tayeh A. The public and domestic domains in the transmission of disease. Trop Med Int Health. 1996;1(1):27–34.

76. Butz WP, Habicht JP, DaVanzo J. Environmental factors in the relationship between breastfeeding and infant mortality: the role of sanitation and water in Malaysia. Am J Epidemiol. 1984;119(4):516–25.

77. Clasen T, Schmidt WP, Rabie T, Roberts I, Cairncross S. Interventions to improve water quality for preventing diarrhoea: systematic review and meta-analysis. BMJ. 2007 Apr 12;334(7597):782.

78. Kawata K. Water and other environmental interventions—the minimum investment concept. Am J Clin Nutr. 1978;31(11):2114–23.

79. Gautam OP, Schmidt WP, Cairncross S, Cavill S, Curtis V. Trial of a Novel Intervention to Improve Multiple Food Hygiene Behaviors in Nepal. Am J Trop Med Hyg. 2017;96(6):1415–26.

80. Kunz R, Oxman AD. The unpredictability paradox: review of empirical comparisons of randomised and non-randomised clinical trials. Bmj. 1998;317(7167):1185–90.

81. Allcott, H. Site Selection Bias in Program Evaluation. Q J Econ. 2015;130(3):1117–65.

82. Esrey SA, Potash JB, Roberts L, Shiff C. Effects of improved water supply and sanitation on ascariasis, diarrhoea, dracunculiasis, hookworm infection, schistosomiasis, and trachoma. Bull World Health Organ. 1991;69(5):609.

83. Clasen TF, Alexander KT, Sinclair D, Boisson S, Peletz R, Chang HH, et al. Interventions to improve water quality for preventing diarrhoea. Cochrane Database Syst Rev. 2015;(10).

84. Hunter PR. Household water treatment in developing countries: comparing different intervention types using meta-regression. Environ Sci Technol. 2009;43(23):8991–7.

85. Wolf J, Prüss-Ustün A, Cumming O, Bartram J, Bonjour S, Cairncross S, et al. Systematic review: assessing the impact of drinking water and sanitation on diarrhoeal disease in low-and middle-income settings: systematic review and meta-regression. Trop Med Int Health. 2014;19(8):928–42.

86. Freeman MC, Stocks ME, Cumming O, Jeandron A, Higgins JP, Wolf J, et al. Systematic review: hygiene and health: systematic review of handwashing practices worldwide and update of health effects. Trop Med Int Health. 2014;19(8):906–16.

87. Wolf J, Hunter PR, Freeman MC, Cumming O, Clasen T, Bartram J, et al. Impact of drinking water, sanitation and handwashing with soap on childhood diarrhoeal disease: updated meta-analysis and meta-regression. Trop Med Int Health. 2018;23(5):508–25.

88. Ross I, Bick S, Ayieko, P, Dreibelbis, R, Allen, E, Freeman, M, et al. Impact of handwashing with soap on acute respiratory infections in low-and middle-income countries – a systematic review [Internet]. PROSPERO CRD42021231414; 2021 [cited 2022 Jul 25]. Available from: https://www.crd.york.ac.uk/prospero/display_record.php?ID=CRD42021231414

89. Morris SS, Black RE, Tomaskovic L. Predicting the distribution of under-five deaths by cause in countries without adequate vital registration systems. Int J Epidemiol. 2003;32(6):1041–51.

90. Benova L, Cumming O, Campbell OM. Systematic review and meta-analysis: association between water and sanitation environment and maternal mortality. Trop Med Int Health. 2014;19(4):368–87.

91. Institute for Resource Development. An assessment of DHS-I data quality. Institute for Resource Development/Macro Systems, Incorporated, Westinghouse, Columbia MD; 1990.

92. Pullum T, Becker S. Evidence of omission and displacement in DHS birth histories: methodological reports vol. 11. Rockv ICF Int. 2014;

93. Schmidt WP, Cairncross S, Barreto ML, Clasen T, Genser B. Recent diarrhoeal illness and risk of lower respiratory infections in children under the age of 5 years. Int J Epidemiol. 2009;38(3):766–72.

94. Pickering AJ, Null C, Winch PJ, Mangwadu G, Arnold BF, Prendergast AJ, et al. The WASH Benefits and SHINE trials: interpretation of WASH intervention effects on linear growth and diarrhoea. Lancet Glob Health. 2019;7(8):e1139–46.

95. Howard G, Bartram J, Brocklehurst C, Colford JM, Costa F, Cunliffe D, et al. COVID-19: urgent actions, critical reflections and future relevance of ‘WaSH’: lessons for the current and future pandemics. J Water Health. 2020;18(5):613–30.

96. Cairncross S, Cliff JL. Water use and health in Mueda, Mozambique. Trans R Soc Trop Med Hyg. 1987 Jan 1;81(1):51–4.

97. Churchill AA, de Ferranti D, Roche R, Tager C, Walters A, Yazer A. Rural water supply and sanitation: time for a change. World Bank; 1987.

98. Moher D. CONSORT: an evolving tool to help improve the quality of reports of randomized controlled trials. Jama. 1998;279(18):1489–91.

99. Schulz KF, Altman DG, Moher D. CONSORT 2010 statement: updated guidelines for reporting parallel group randomised trials. J Pharmacol Pharmacother. 2010;1(2):100–7.

100. Bose R. A checklist for the reporting of randomized control trials of social and economic policy interventions in developing countries: CEDE Version 1.0. New Delhi: International Initiative for Impact Evaluation; 2010.

101. Kremer M, Luby S, Maertens R, Tan B. Water treatment and child mortality: a systematic review and meta-analysis [Internet]. [cited 2022 Jul 26]. Available from: Available at: https://cpb-us-w2.wpmucdn.com/voices.uchicago.edu/dist/0/2830/files/2022/02/Water-meta-analysis-manuscript-2022.02.20.docx.pdf

102. Gallandat K, Cumming O. Impact Evaluation of Urban Water Supply Improvements on Cholera and Other Diarrhoeal in Uvira, Democratic Republic of Congo [Internet]. clinicaltrials.gov; 2022 Aug [cited 2022 Sep 15]. Report No.: NCT02928341. Available from: https://clinicaltrials.gov/ct2/show/NCT02928341

103. Cha S, Kang D, Tuffuor B, Lee G, Cho J, Chung J, et al. The Effect of Improved Water Supply on Diarrhea Prevalence of Children under Five in the Volta Region of Ghana: A Cluster-Randomized Controlled Trial. Int J Environ Res Public Health. 2015 Oct;12(10):12127–43.

104. Devoto F, Duflo E, Dupas P, Parienté W, Pons V. Happiness on tap: Piped water adoption in urban Morocco. Am Econ J Econ Policy. 2012;4(4):68–99.

105. Waddington HS, Villar PF, Valentine JC. Can Non-Randomised Studies of Interventions Provide Unbiased Effect Estimates? A Systematic Review of Internal Replication Studies. Eval Rev. 2022;0193841X221116721.

